# Genetics of Low Polygenic Risk Score Type 1 Diabetes Patients: rare variants in 22 novel loci

**DOI:** 10.1101/2020.10.13.20211987

**Authors:** Jingchun Qu, Hui-Qi Qu, Jonathan Bradfield, Joseph Glessner, Xiao Chang, Lifeng Tian, Michael March, Jeffrey D Roizen, Patrick Sleiman, Hakon Hakonarson

## Abstract

With polygenic risk score (PRS) for autoimmune type 1 diabetes (T1D), this study identified T1D cases with low T1D PRS and searched for susceptibility loci in these cases. Our hypothesis is that genetic effects (likely mediated by relatively rare genetic variants) of non-mainstream (or non-autoimmune) T1D might have been diluted in the previous studies on T1D cases in general. Two cohorts for the PRS modeling and testing respectively were included. The first cohort consisted of 3,356 T1D cases and 6,203 controls, and the independent second cohort consisted of 3,355 T1D cases and 6,203 controls. Cases with low T1D PRS were identified using PRSice-2 and compared to controls with low T1D PRS by genome-wide association (GWA) test. Twenty-six genetic loci with SNPs/SNVs associated with low PRS T1D at genome-wide significance (P≤5.0xE-08) were identified, including 4 established T1D loci, as well as 22 novel loci represented by rare SNVs. For the 22 novel loci, 12 regions have been reported of association with obesity related traits by previous GWA studies. Five loci encoding long intergenic non-protein coding RNAs (lncRNA), two loci involved in N-linked glycosylation, two loci encoding GTPase activators, and two ciliopathy genes, are also highlighted in this study.

## Introduction

Type 1 diabetes (T1D) is caused by T-cell mediated autoimmune destruction of pancreatic β-cells(1). There is no cure for T1D to date. The molecular mechanisms underlying T1D are complex and not completely understood. Human genetic studies have uncovered multiple T1D genes that contribute to our understanding of the pathogenesis ofT1D(2-7). With the rapid advances in human genomics technology in recent years, over 70 T1D loci have been identified(8) (https://www.ebi.ac.uk/gwas/). While these discoveries of T1D-associated genes have greatly increased our knowledge of T1D, our current genetic knowledge on T1D is far from complete, and a large number of T1D genes remain uncovered(9). A key bottleneck for the GWAS approach is limitation of sample size even with the presense of collaborative international consortia(10). The phenotype of type 1 diabetes has been regarded as heterogeneous. While the majority of T1D patients have autoimmune disease, 5–10% of Caucasian diabetic subjects with recent-onset T1D do not have islet cell antibodies, often referred to as T1bD(11). Due to different pathogenesis, T1bD cases may be associated with different genetic loci from autoimmune T1D, or T1aD. However, the smaller proportion of T1bD cases suggests that T1bD-related genetic effects have been diluted in the previous studies with T1D cases studied in general. Besides T1bD, the non-autoimmune and monogenic form of pediatric diabetes, maturity-onset diabetes of the young (MODY) cases, may be misdiagnosed as T1D(12), which further contributes to the heterogeneity of the T1D phenotype.

With numerous genetic loci for many human complex diseases identified to date, polygenic risk scores (PRS) aggregate the effects of many genetic variants across the human genome into a single score, an approach that has been shown of improve disease prediction and differential diagnosis(13). The T1D loci identified by the GWAS studies to date are mainly associated with the genetic susceptibility of the major component of the heterogeneous T1D phenotype, i.e. T1aD, while the genetic susceptibility of the minor non-autoimmune components (e.g. T1bD and misdiagnosed MODY) are undere-represented in those results likely as a result of being diluted In this study, we propose that a high T1D PRS score predicts or suggests a T1aD case, whereas a low T1D PRS score in a T1D case suggests the opposite and represents our major interest in this study. Our aim in this study is to identify low PRS T1D cases and to run a seprate GWAS in an attempt to uncover genetic loci associated with T1bD patients.

## Methods

Subjects: 6,711 European T1D cases and 12,406 European controls were included in this study. The T1D cases were from the Children’s Hospital of Philadelphia (CHOP)(14), The Diabetes Control and Complications Trial – Epidemiology of Diabetes Interventions and Complications (DCCT-EDIC) cohort (http://www.ncbi.nlm.nih.gov/projects/gap/cgi-bin/study.cgi?study_id=phs000086.v2.p1), the Type 1 Diabetes Genetics Consortium (T1DGC, http://www.ncbi.nlm.nih.gov/projects/gap/cgi-bin/study.cgi?study_id=phs000180.v1.p1), and later recruited subjects at CHOP, respectively. The genotyping was done by the Illumina Human Hap550 Genotyping BeadChip or a newer version of Illumina Genotyping BeadChip. Other demographic, phenotypic and genotypic details about these individuals were described in our previous publication(15). Imputation of 39,131,579 single nucleotide polymorphisms (SNP) on auto-chromosomes was done using the Sanger Imputation Service (https://www.sanger.ac.uk/tool/sanger-imputation-service/) based on the Haplotype Reference Consortium (HRC) r1.1 reference panel (HRC.r1-1.GRCh37.wgs.mac5.sites.tab), with the quality filters of R^2^ ≥ 0.4. Altogether, 32,251,301 autosomal single nucleotide variants (SNV) with quality R^2^ ≥ 0.4 were included in this study. Population stratification was assessed by principal component analysis (PCA), and genetic association tests were corrected by the first 10 principal components (PC). The association test was done using PLINK1.9 software(16).

Polygenic risk scores (PRS): To avoid the issue of overfitting for PRS scoring, the subjects were randomly splitted into two independent cohorts without duplication, i.e. the PRS training cohort including 3,356 T1D cases and 6,203 controls, and the PRS testing cohort including 3,355 T1D cases and 6,203 controls. PRSs of the test cohort were calculated using the Polygenic Risk Score software (PRSice-2)(17), based on the statistics of the training group. The performance of a series of cutoff of T1D association P-values (including 10^−10^, 10^−9^, 10^−8^, 10^−7^, 10^−6^, 10^−5^, 10^−4^, 0.001, 0.01, 0.05, 0.1, 0.2, and 1) for selection of SNP markers was assessed by the Area Under the ROC Curve (AUC). The P-value cutoff with the largest AUC was adopted.

GWAS of T1D patients with low PRS: According to the PRS values, the T1D patients were separated into two groups, i.e. a low PRS group and a high PRS group. The PRS cutoff was determined by the maximum Matthews correlation coefficient (MCC). Using the same PRS cutoff, health controls with low T1D PRS were identified. The GWAS of T1D patients with low PRS was performed by comparing to health controls with low T1D PRS. The Manhattan plots were done using the web-based FUMA platform(18). Genetic association signals within each locus were plotted by LocusZoom(19).

### Data and Resource Availability

The datasets generated during and/or analyzed during the current study are available from the corresponding author upon reasonable request.

## Results

### AUC of different cutoffs of T1D association P-values for SNP selection and PRS

The AUCs of different cutoffs of T1D association P-values for selection of SNP sets are shown in Table 1a. The best AUC (0.8607) is seen at the cutoff of P-value≤1E-05, which suggests that stricter cutoff may cause the missing of informative SNPs, while looser may introduce noise by including SNPs with spurious T1D association. Based on the SNP markers with T1D association P-value≤1E-05, a PRS score was acquired for each individual in the independent test cohort. By the maximum MCC (Supplementary Table 1), a PRS cutoff of 1.11E-03 has the maximum MCC (0.6294). A PRS≤1.11E-03 was defined as low risk, and a PRS>1.11E-03 was defined as high risk. With this threshold, the sensitivity (True positive rate, TPR) for T1D prediction is 75.9%, and the specificity (True negative rate, TFR) for T1D prediction is 86.4%. By PRS≤1.11E-03, 810 (24.1%, including 408 males, 400 females, and 2 cases with undetermined sex) out of 3,355 T1D cases had low PRS; and 5,358 (86.4%, including 2,893 males, 2,453 females, and 12 cases with undetermined sex) out of 6,203 controls had low PRS.

**Table 1.** The AUCs of different cutoffs of T1D association P-values

| **a. First cohort** |  |
| --- | --- |
| **P value\*** | **AUC\*\*** |
| ≤1.00E-10 | 0.8462 |
| ≤1.00E-09 | 0.8487 |
| ≤1.00E-08 | 0.8518 |
| ≤1.00E-07 | 0.8565 |
| ≤1.00E-06 | 0.8604 |
| ≤1.00E-05 | 0.8607 |
| ≤1.00E-04 | 0.8590 |
| ≤0.001 | 0.8561 |
| ≤0.01 | 0.8546 |
| ≤0.05 | 0.8502 |
| ≤0.1 | 0.8508 |
| ≤0.2 | 0.8530 |
| ≤0.5 | 0.8563 |
| ≤1 | 0.8579 |
| **b. Switched cohort** |  |
| **P value\*** | **AUC\*\*** |
| ≤1.00E-10 | 0.8576 |
| ≤1.00E-09 | 0.8589 |
| ≤1.00E-08 | 0.8588 |
| ≤1.00E-07 | 0.8609 |
| ≤1.00E-06 | 0.8633 |
| ≤1.00E-05 | 0.8654 |
| ≤1.00E-04 | 0.8618 |
| ≤0.001 | 0.8555 |
| ≤0.01 | 0.8470 |
| ≤0.05 | 0.8441 |
| ≤0.1 | 0.8446 |
| ≤0.2 | 0.8467 |
| ≤0.5 | 0.8521 |
| ≤1 | 0.8533 |

\* The P values are based on the statistics of the PRS training cohort;
\*\* The AUCs are the PRS performances in the independent testing cohort.

### GWAS of T1D patients with low PRS

The GWAS of T1D patients with low T1D PRS compared to controls with low T1D PRS identified a large number of SNPs associated with T1D with genome-wide significance (P≤5.0xE-08), from 7 genetic loci (Supplementary Table 2, Figure 1). Among these 7 genetic loci, 3 loci have been established of T1D association by previous studies, including *HLA, INS*, and *PTPN22* (Table 2a). By looking at the established leading T1D signal of each locus, the frequencies of the predisposing alleles of *HLA* and *PTPN22* were lower in the low T1D PRS cohort, while the protective allele of *INS* were higher in the low T1D PRS cohort. The effect sizes of *HLA* (P=2.72E-06) and *PTPN22* (P=0.047) were significantly smaller in the low PRS cases. Besides these 3 established T1D loci, 4 novel loci associated with low PRS T1D were identified (Table 3a). LocusZoom plots for genetic association signals within each locus are shown in Supplementary Figure 1-4. The association signals of these loci are only seen in low PRS T1D cases, but not in the T1D cases overall, and were missed previously due to diluted genetic effects.

**Table 2.**
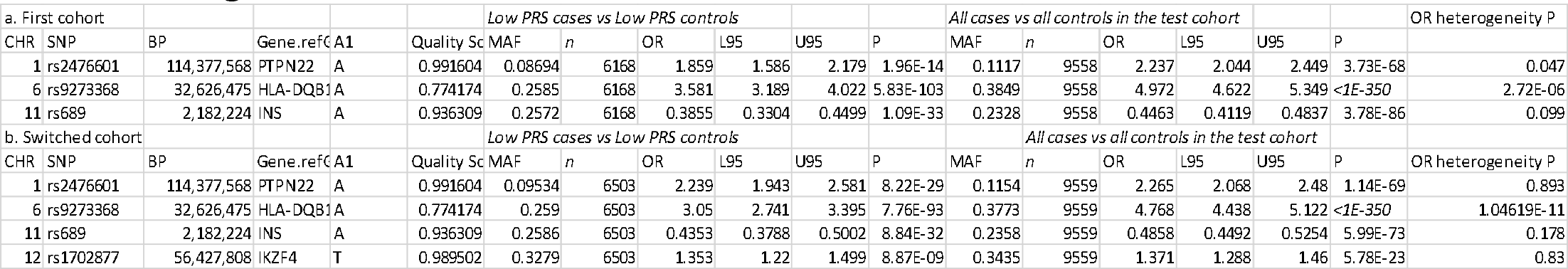
Leading SNPs at three loci have been established of T1D association

**Table 3.**
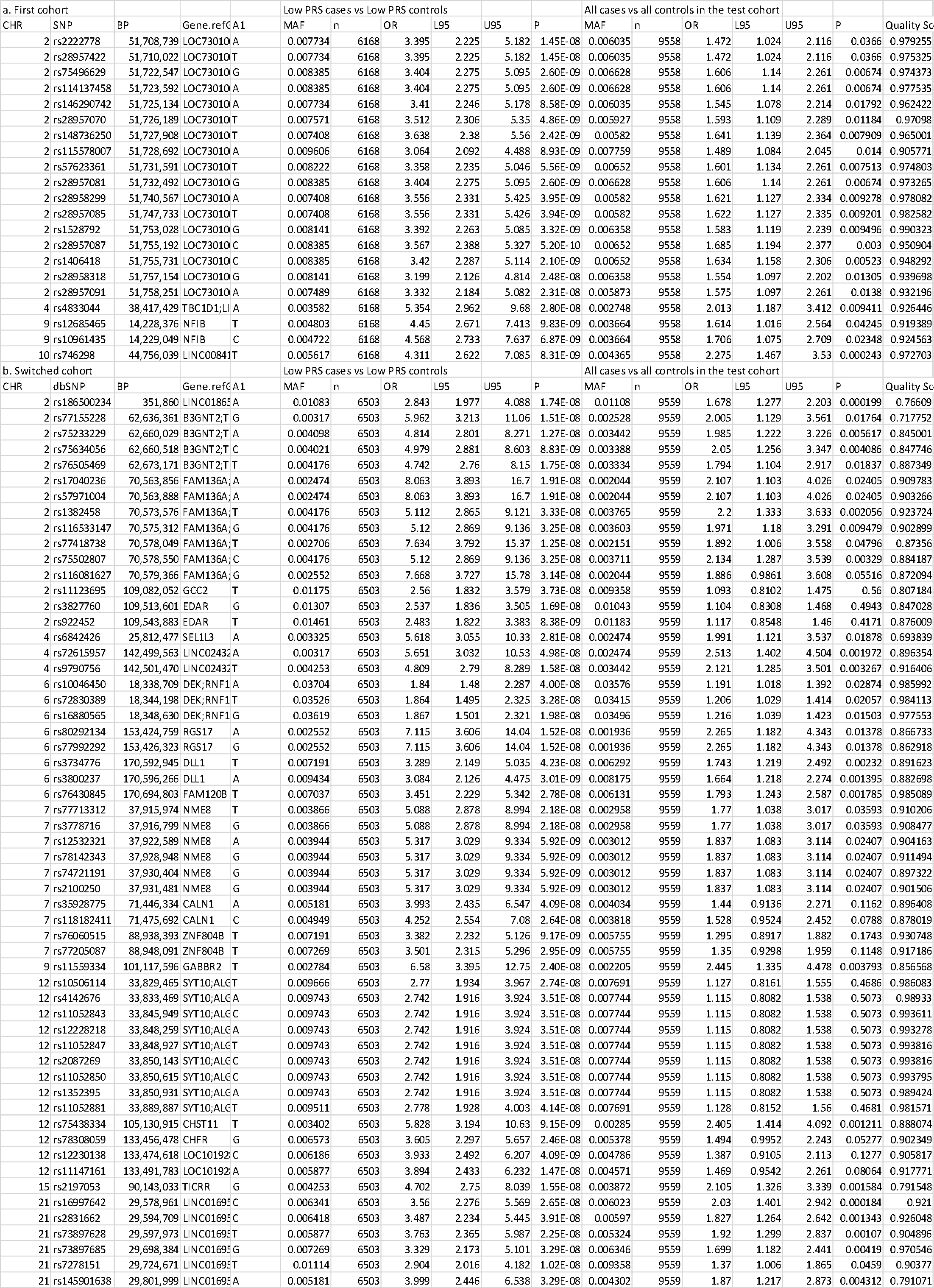
Novel loci associated with low PRS T1D

**Figure 1.**
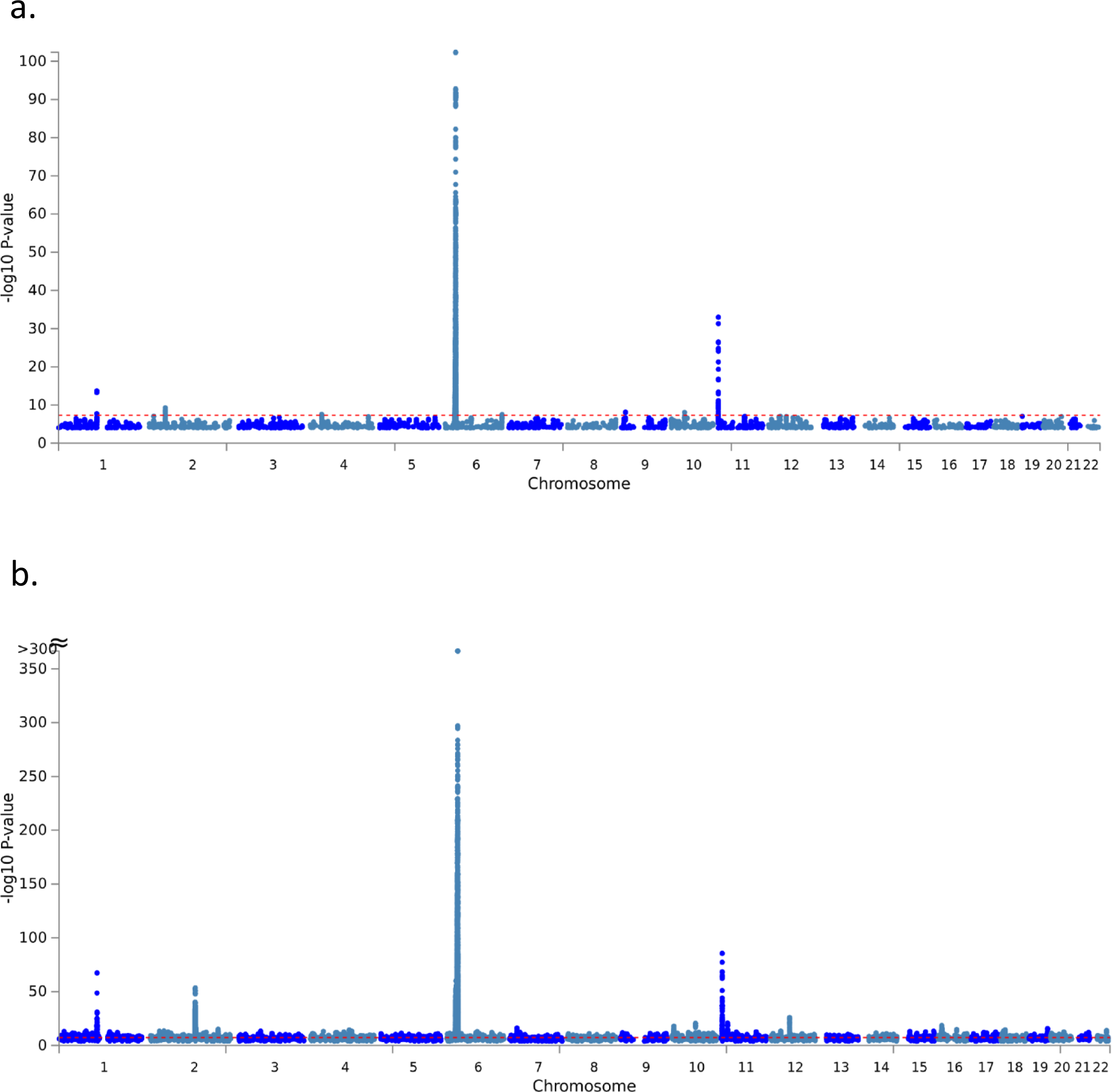
The Manhattan plots of the first cohort. (a) The plot of the GWAS of T1D patients with low T1D PRS compared to controls with low T1D PRS (810 cases vs. 5358 controls); (b) The plot of the GWAS of all T1D patients compared to all controls (3355 cases vs. 6203 controls).

### Replication of the PRS model and additional novel loci

Consequently, we switched the two cohorts, i.e. using the second cohort for the statistics of PRS modelling, then we tested the PRS models in the first cohort. The AUCs of different cutoffs of T1D association P-values for selection of SNP sets are shown in Table 1b. The best AUC (0.8654) is seen at the cutoff of P-value≤1E-05, which repeated the PRS model in the above step. Based on the SNP markers with T1D association P-value≤1E-05, a PRS score was acquired for each individual in the independent test cohort. By the maximum MCC (Supplementary Table 3), a PRS cutoff of 1.24E-03 has the maximum MCC (0.6294). A PRS≤7.18E-04 was defined as low risk, and a PRS>7.18E-04 was defined as high risk. With this threshold, the sensitivity (True positive rate, TPR) for T1D prediction is 66.0%, and the specificity (True negative rate, TFR) for T1D prediction is 93.6%. By PRS≤7.18E-04, 918 (27.4%, including 437 males, 479 females, and 2 cases with undetermined sex) out of 3,356 T1D cases had low PRS; and 5,585 (90.0%, including 3,008 males, 2,565 females, and 12 cases with undetermined sex) out of 6,203 controls had low PRS.

As expected from the above results, in the switched cohort, the GWAS of T1D patients with low T1D PRS compared to controls with low T1D PRS identified a large number of SNPs associated with T1D with genome-wide significance (P≤5.0xE-08) as well (Supplementary Table 4, Figure 2). Among these loci, 4 loci have been established of T1D association by previous studies, including *HLA, INS, PTPN22*, and the *IKZF4/RPS26/ERBB3* locus (Table 2b). Consistent to the first GWAS results listed above, by looking at the established leading T1D signal of each locus, the frequencies of the predisposing alleles of *HLA, PTPN22* and *IKZF4* were lower in the low T1D PRS cohort, while the protective allele of *INS* were higher in the low T1D PRS cohort. The effect size of the leading *HLA* SNP was significantly smaller in the low PRS cases (P=1.05E-11). Besides these established T1D loci, 18 novel loci associated with low PRS T1D were identified in this cohort (Table 3b). LocusZoom plots for genetic association signals within each locus are shown in Supplementary Figure 5-22.

**Figure 2.**
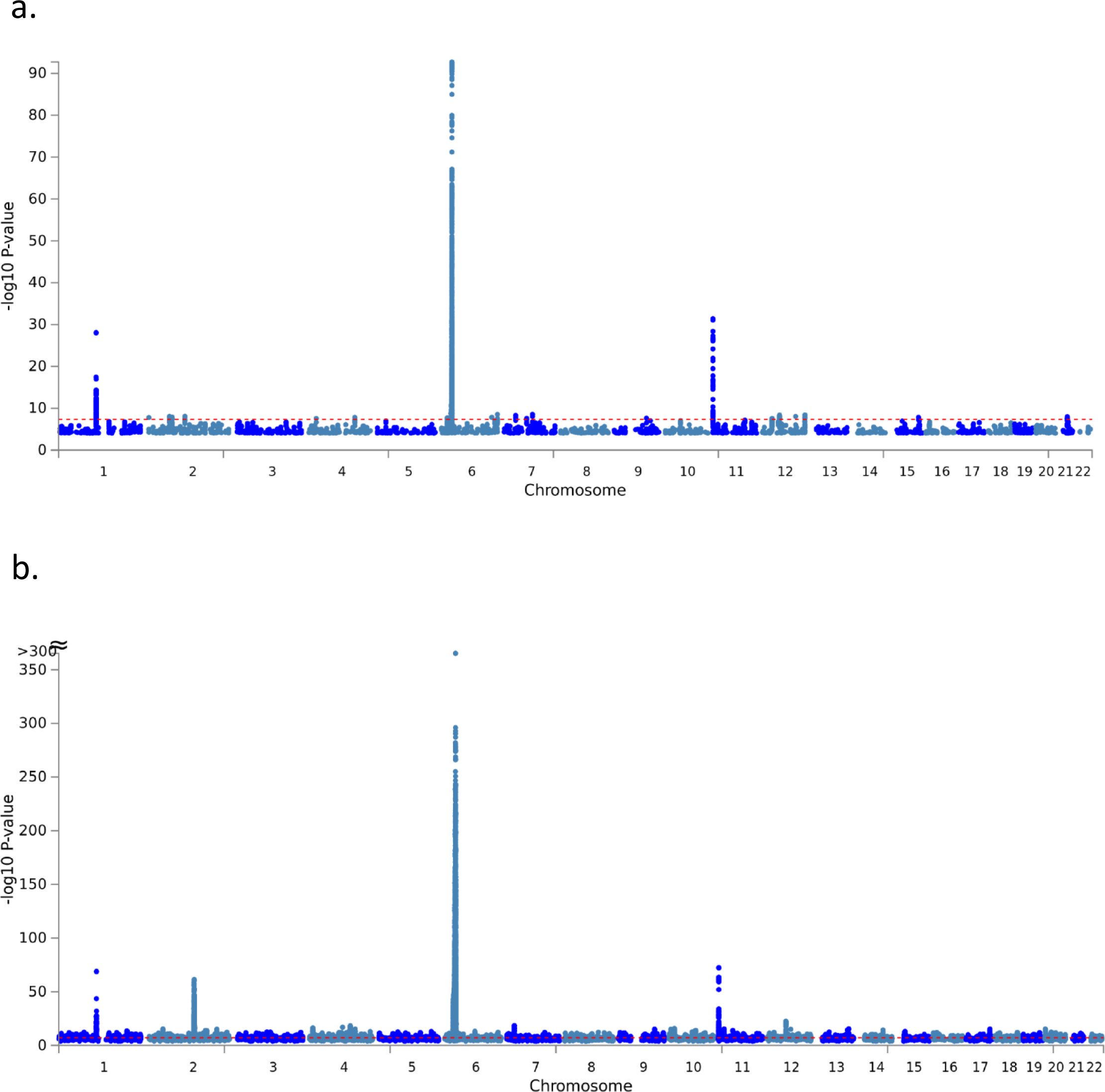
The Manhattan plots of the second cohort. (a) The plot of the GWAS of T1D patients with low T1D PRS compared to controls with low T1D PRS (918 cases vs. 5585 controls); (b) The plot of the GWAS of all T1D patients compared to all controls (3356 cases vs. 6203 controls).

## Discussion

Altogether, rare variants (MAF<5%) from 22 novel loci were identified in the low PRS T1D cases with genome-wide significance (P<5.00E-08), in addition to the 4 established T1D loci with smaller genetic effects in these cases. The association signals of these loci are only seen in low PRS T1D cases, but not in the T1D cases overall, and were missed previously due to rare allele frequencies and diluted genetic effects in the general T1D cohort. Among the 22 loci, two genetic regions have been reported of association with diabetes, i.e. the region containing the *DLL1/ FAM120B* locus associated with type 1 diabetes in Caucasian by our previous study(20), and the region containing the *TICRR* locus associated Type 2 diabetes in African population(21). In addition, a number of genetic associations with body mass index (BMI), obesity, and autoimmunity, have been reported in the flanking regions of 300kb on each side of the new loci according to the GWAS Catalog (https://www.ebi.ac.uk/gwas/, Supplementary materials). Further details on these 22 loci are described below.

### *LINC01865/LINC01874* tagged by rs186500234

The long intergenic non-protein coding RNA 1865 gene (*LINC01865*) has low expression observed in testis, brain, and duodenum. The long intergenic non-protein coding RNA 1874 gene (*LINC01874*) has restricted expression toward kidney(22). This genetic region has been reported of association with body mass index (BMI) by previous study(23).

### *LOC730100* tagged by rs28957087

*LOC730100* encodes a long non-coding RNA (ncRNA), a competing endogenous RNA for human microRNA 760 (miR-760)(24). The latter inhibits the expression of the Forkhead Box A1 gene (*FOXA1*). As a hepatocyte nuclear factor, *FOXA1*, also known as *HNF3A* or *TCF3A*, regulates tissue-specific gene expression in liver and many other tissues(25). FoxA1 is essential for normal pancreatic and ß-cell function and a negative regulator of the hepatocyte nuclear factor-1 (HNF1) homeobox A gene (*HNF1A*) and the hepatocyte nuclear factor 4, alpha gene (*HNF4A*)(26) (27). *HNF1A* and *HNF4A* are established genes causing maturity-onset diabetes of the young (MODY). The *FOXA1* mutation Ser448Asn has been suggested of association with impaired glucose homeostasis(27).

### *B3GNT2/TMEM17* tagged by rs75634056

The UDP-GlcNAc:betaGal beta-1,3-N-acetylglucosaminyltransferase 2 gene (*B3GNT2*) encodes an enzyme involved in the biosynthesis of poly-N-acetyllactosamine chains. The gene plays an important role in immunological biofunctions, and its deficiency causes hyperactivation of lymphocytes in mice(28). The transmembrane protein 17 gene (*TMEM17*) encodes a critical component of a protein complex at the base of cilia. Previous GWAS studies have reported association with Crohn’s disease, ankylosing spondylitis, and hypothyroidism in this genetic region.

### *FAM136A/TGFA* tagged by rs77418738

The family with sequence similarity 136 member A gene (*FAM136A*) encodes a mitochondrially localized protein. The transforming growth factor alpha gene (TGFA) mediates cell-cell adhesion and activates cell proliferation, differentiation and development. This region has been reported of association with obesity-related traits(29).

### *GCC2/EDAR* tagged by rs922452

The GRIP and coiled-coil domain containing 2 (GCC2) encodes a long coiled-coil protein, also known as GCC185, which is localized to the trans-Golgi network with critical function in maintaining Golgi structure and tethering transport vesicle(30). The ectodysplasin A receptor gene (EDAR) encodes a member of the tumor necrosis factor receptor family with a key role in ectodermal differentiation. Association with low birth weight at this region has been reported(31).

### *SEL1L3* tagged by rs6842426

The locus SEL1L family member 3 gene (*SEL1L3*) is a paralog of the SEL1L adaptor subunit of ERAD E3 ubiquitin ligase gene (*SEL1L*). The latter is highly expressed in pancreas and thyroid, and is crucial for misfolded proteins in the endoplasmic reticulum being discharged into the cytosol and degraded by the proteasome(32). This gene region has been reported association with obesity-related traits(29) and non-alcoholic fatty liver disease(33).

### *TBC1D1/LINC01258* tagged by rs4833044

This genetic locus contains two genes, the TBC1 domain family member 1 gene (*TBC1D1*) and the long intergenic non-protein coding RNA 1258 gene (*LINC01258*). A common variant it this locus has been reported to be associated with childhood obesity(29; 34), triacylglycerol 54:5 levels(35), lymphocyte percentage of leukocytes(36) by previous studies. Acting as a GTPase activator, the *TBC1D1* protein plays a role in regulating cell growth and differentiation. Rare mutations in *TBC1D1* have been reported to be associated with congenital anomalies of the kidney and urinary tract.

### *LINC02432/IL15* tagged by rs9790756

The long intergenic non-protein coding RNA 2432 gene (*LINC02432*) has higher expression in kidney and pancreas. Interleukin 15 (IL-15) encoded by the gene *IL15* is essential for regulating activation and proliferation of T and natural killer cells, and supporting lymphoid homeostasis. IL-15 and interleukine 2 (IL-2) share many biological activities and receptor components with IL-2. IL-2 is a powerful growth factor for both T and B lymphocytes. Both IL2 and the α chain of the IL2 receptor complex gene (*IL2RA*) has been established of genetic association with T1D by previous studies(37-39).

### *DEK/RNF144B* tagged by rs16880565

The DEK proto-oncogene gene (*DEK*) encodes a site-specific DNA binding protein and a component of the pre-mRNA splicing complex, and is involved in transcriptional regulation and pre-mRNA splicing. *DEK* encoded protein is also an autoantigen in patients with pauciarticular onset juvenile rheumatoid arthritis. The ring finger protein 144B gene (*RNF144B*) encoded protein inhibits LPS-induced inflammatory responses by binding with TANK binding kinase 1 (TBK1) and causing interferon regulatory factor 3 (IRF3) dephosphorylation and interferon β (IFN-β) reduction. This region has been reported of association with BMI by previous studies(23; 40).

### *RGS17* tagged by rs80292134

The regulator of G protein signaling 17 gene (*RGS17*) encodes a member of the regulator of G-protein signaling family. This genetic region has been established association with BMI by previous studies(23; 40).

### *DLL1*/*FAM120B* tagged by rs3800237

The delta like canonical Notch ligand 1 (DLL1) encodes a Notch ligand with a role in cell-fate decision processes in lymphopoiesis. This Notch ligand can completely inhibit the differentiation of human hematopoietic progenitors into the B cell lineage while promoting the generation of T cell/natural killer (NK) precursors(41). The family with sequence similarity 120B gene (FAM120B) encodes a constitutive coactivator of peroxisome proliferator-activated receptor γ (PPARγ, a major therapeutic target for insulin sensitivity) and promotes adipogenesis(42). The region containing the *DLL1/ FAM120B* genes has been reported of association with T1D in Caucasian by our previous study(20).

### *NME8/GPR141* tagged by rs12532321

The NME/NM23 family member 8 gene (*NME8*) encodes an axoneme protein, and its mutation may cause primary ciliary dyskinesia. The G protein-coupled receptor 141 gene (*GPR141*) at the upstream of *NME8* is highly expressed in bone marrow. This genetic region has been reported of association with obesity-related traits in Hispanic children(29).

### *CALN1* tagged by rs118182411

The calneuron 1 gene (*CALN1*), encoding a protein with high similarity to the calcium-binding proteins of calmodulin, is highly expressed in brain and adrenal. This genetic region has established association with BMI by previous studies(23; 40).

### *ZNF804B* tagged by rs77205087

The zinc finger protein 804B gene (*ZNF804B*) has been reported of association with N-linked glycosylation of human immunoglobulin G (IgG), which modulates its binding to Fc receptors(43). N-glycosylation of cytokines and proteases is also a regulatory mechanism in inflammation and autoimmunity(44). Changes in N-glycosylation have been associated with different autoimmune diseases, including rheumatoid arthritis(45), type 1 diabetes(46), Crohn’s disease(47).

### *NFIB* tagged by rs10961435

The nuclear factor I B gene (*NFIB*) encodes a transcription factor in the FOXA1 transcription factor network. NFIB has been shown to play critical roles in lung and brain development. A previous study has shown that NFIB can bind with FoxA1 and modulate the transcriptional activity of FoxA1(48), while the later has been suggested to play a role in pancreatic and ß-cell function and non-autoimmune diabetes as discussed above.

### *TBC1D2/GABBR2* tagged by rs11559334

This genetic locus contains two protein-coding genes, the TBC1 domain family member 2 gene (*TBC1D2*) and the gamma-aminobutyric acid type B receptor subunit 2 gene (*GABBR2*, encoding a member of the G-protein coupled receptor 3 family). As discussed above, this study identified an association signal in the *TBC1D1* region, and the *TBC1D1* locus has been reported of association with childhood obesity(29; 34).

### *LINC00841/C10orf142* tagged by rs746298

The two genes at this locus, *LINC00841/C10orf142*, encode two long intergenic non-protein coding RNAs (lincRNA). While the function of these two genes remain unknown, this locus has been reported of association with obesity-related traits(29).

### *SYT10/ALG10* tagged by rs10506114

The synaptotagmin 10 gene (*SYT10*) encodes a membrane protein of secretory vesicles expressed in pancreas, lung and kidney(49). The ALG10 alpha-1,2-glucosyltransferase gene (*ALG10*) encodes a membrane-associated protein that adds the third glucose residue to the lipid-linked oligosaccharide precursor for N-glycosylation in endoplasmic reticulum (ER)(50). As discussed above in the *ZNF804B* locus, N-glycosylation of IgG, cytokines and proteases is also a regulatory mechanism in inflammation and autoimmunity(43; 44) associated with different autoimmune diseases. This region has established association with waist-hip ratio by previous study(40).

### *CHST11* tagged by rs75438334

The carbohydrate sulfotransferase 11 gene (*CHST11*) encodes a member of the sulfotransferase 2 family catalyzing chondroitin sulfate synthesis. This genetic region has been reported of association with waist circumference adjusted for body mass index by previous study(51).

### *CHFR/LOC101928530/ZNF605* tagged by rs12230138

The checkpoint with forkhead and ring finger domains gene (*CHFR*) encodes an E3 ubiquitin-protein ligase and is involved in the DNA damage response and checkpoint regulation. The structure and function of the gene *LOC101928530* is still uncharacterized. The function of the zinc finger protein 605 gene (*ZNF605*) may be related to Herpes Simplex Virus 1 infection (https://pathcards.genecards.org/card/herpes_simplex_virus_1_infection). This region has been reported of association with BMI by previous study(52).

### *TICRR/ KIF7* tagged by rs2197053

The TOPBP1 interacting checkpoint and replication regulator gene (*TICRR*) encodes Treslin, which is involved in triggering the initiation of DNA replication. The kinesin family member 7 gene (*KIF7*) in this region encodes a cilia-associated protein of the kinesin family, with its mutations causing ciliopathies. The region containing the *TICRR* gene has been reported of association with T2D in African population(21), BMI(23; 52), and obesity-related traits(29) by previous studies.

### *LINC01695/LINC00161* tagged by rs7278151

Function of the long intergenic non-protein coding RNA 1695 gene (*LINC01695*) is still uncharacterized. The long intergenic non-protein coding RNA 161 gene (*LINC00161*) encodes a functional RNA that regulates Mitogen-activated protein kinase 1 (MAPK1) expression. The MAPK1/STAT3 pathway has been proposed as a novel diabetes target for its critical role in glucose homeostasis(53).

In summary, in the genetic regions containing the 22 novel loci disclosed by this study, more than half of these regions have been reported of association with obesity-related traits, BMI, or waist circumference. The correlation with obesity related traits or impaired glucose homeostasis is in keeping with non-autoimmune roles in the diabetes patients with low T1D PRS. Interestingly, genes related N-linked glycosylation, e.g. *ZNF804B* and *ALG10*, are highlighted in this study, which may suggest the role of N-glycosylation bridging impaired glucose homeostasis and autoimmune diabetes. N-glycosylation is commonly altered in diabetes(54). This particular locus supports an interesting hypothesis of T1D pathogenesis, i.e. the accelerator hypothesis, which implies that increasing obesity-associated insulin resistance accelerates the disease process of type 1 diabetes(55; 56). Insulin resistance-related mechanisms might thus be able to serve as potential novel therapeutic targets for these patients with low T1D PRS.

In addition, 5 loci encoding long intergenic non-protein coding RNAs (lncRNA) identified in this study emphasize the importance of lncRNAs in these diabetes patients. This study identified 2 loci containing *TBC1D1* and *TBC1D2* respectively, encoding two GTPase activators. *TBC1D1* has been suggested as a novel obesity gene by previous study(34). Two loci containing the *TMEM17* and *KIF7* genes corrected with ciliopathies suggest a role of primary cilia in diabetes(57). However, we admit that this study has limitations related to the bottleneck of sample size and data resources. The novel loci reported in this study still need replication in independent samples. In addition, the functional mechanisms of these genetic loci in diabetes warrant experimental investigation.

## Acknowledgement

The authors apologize that many important references listed in the supplementary materials cannot be cited in the main text because of page limitation. The study was supported by Institutional Development Funds from the Children’s Hospital of Philadelphia to the Center for Applied Genomics and The Children’s Hospital of Philadelphia Endowed Chair in Genomic Research to HH. Dr. Hakon Hakonarson is the guarantor of this work and, as such, had full access to all the data in the study and takes responsibility for the integrity of the data and the accuracy of the data analysis.

## Competing interests

none to declare.

## Notes

### Competing Interest Statement

The authors have declared no competing interest.

### Funding Statement

The study was supported by Institutional Development Funds from the Childrens Hospital of Philadelphia to the Center for Applied Genomics and The Childrens Hospital of Philadelphia Endowed Chair in Genomic Research to HH.

### Author Declarations

The study was approved by the institutional review board and the ethics committee of the Children's Hospital of Philadelphia.

