## Supplementary figure for "Genetics of Low Polygenic Risk Score Type 1 Diabetes Patients: rare variants in 22 novel loci"

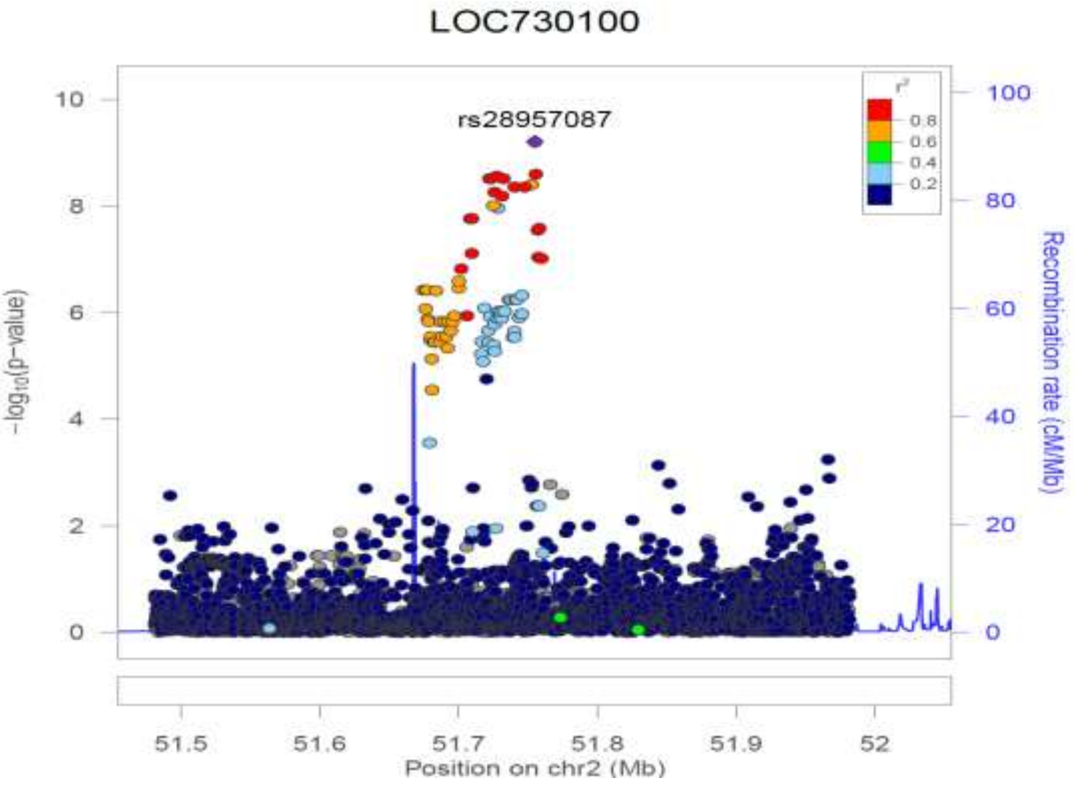


a.

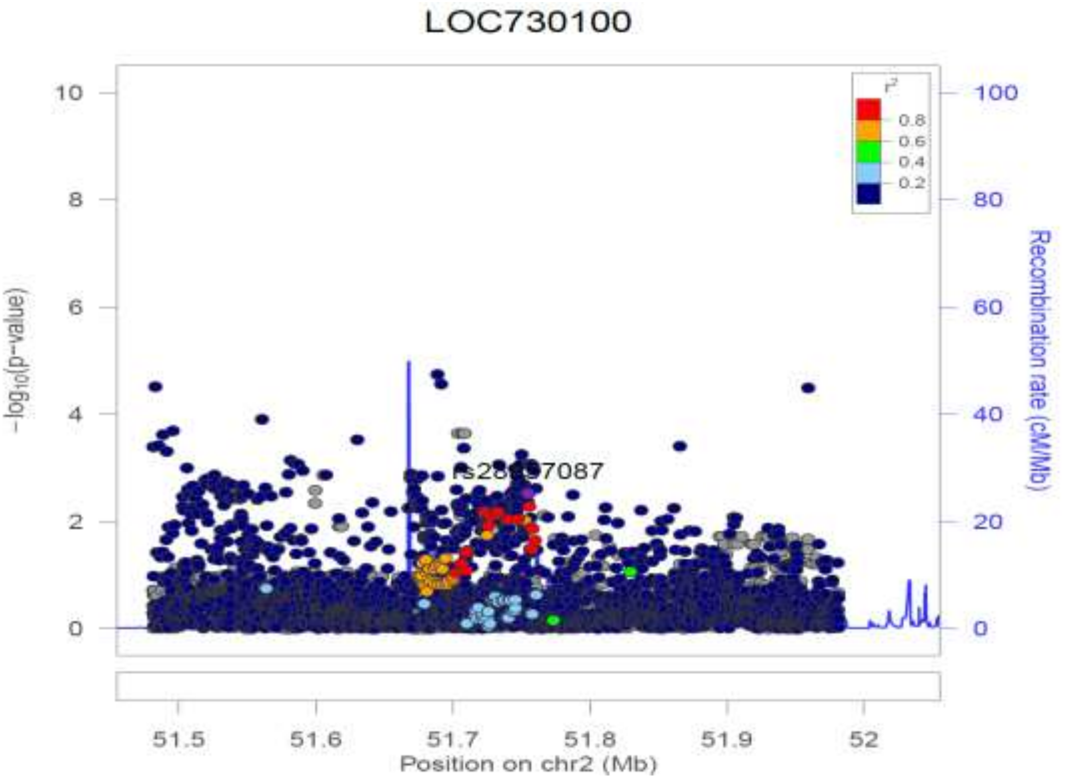


b.

**Supplementary Figure 1.** The LocusZoom plots for each of the *LOC730100* locus. (a) The plot of the association tests of T1D patients with low T1D PRS compared to controls with low T1D PRS; (b) The plot of the association tests of all T1D patients compared to all controls.

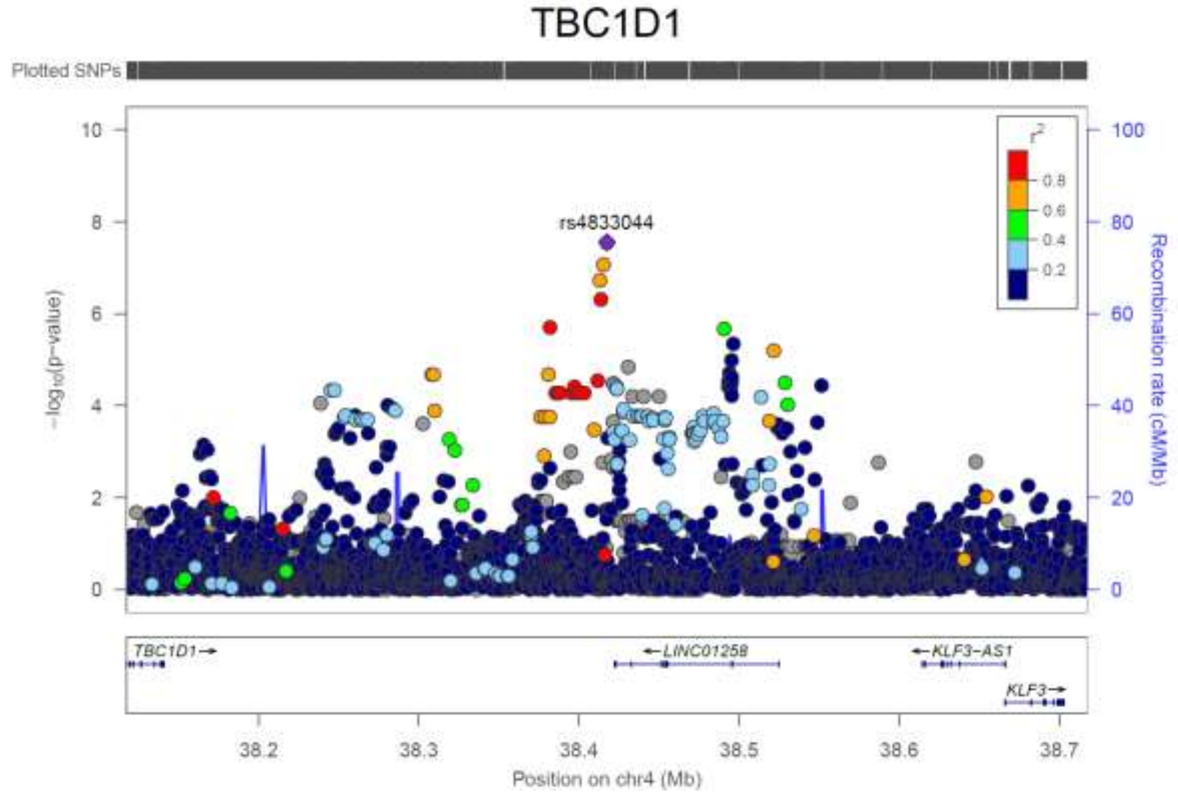


a.

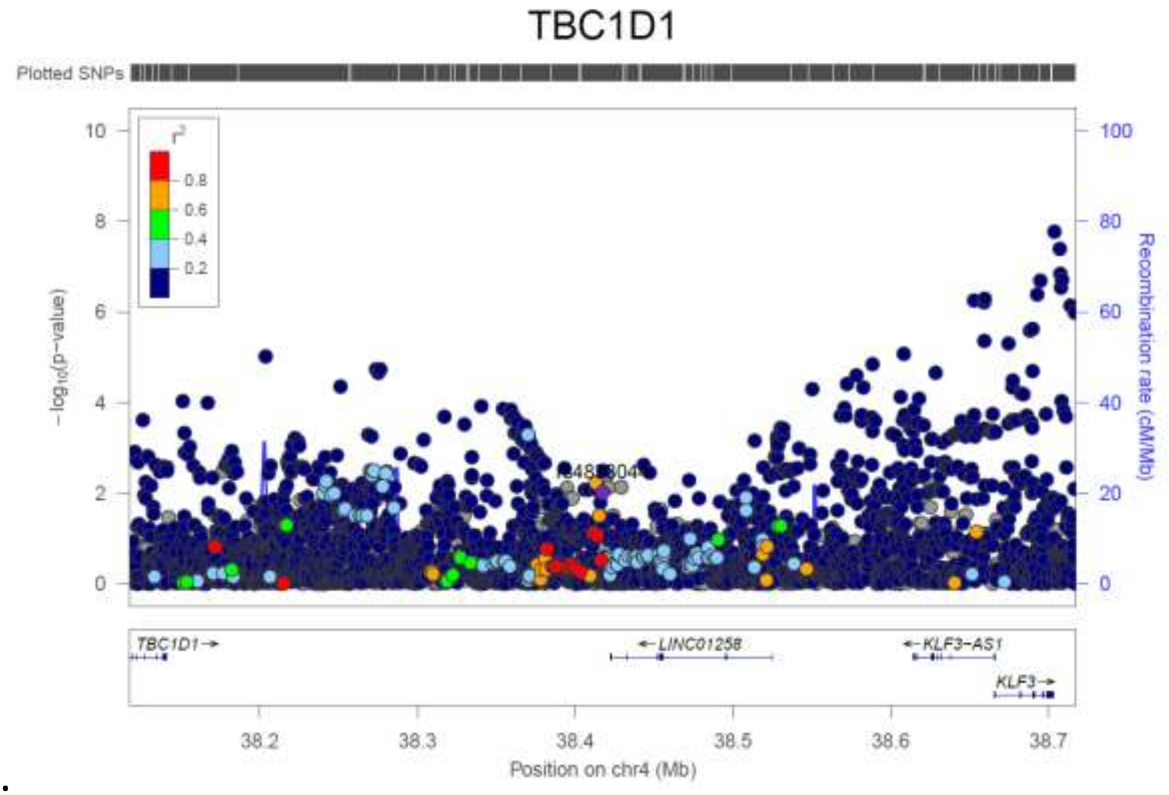


b.

**Supplementary Figure 2.** The LocusZoom plots for each of the *TBC1D1/LINC01258* locus. (a) The plot of the association tests of T1D patients with low T1D PRS compared to controls with low T1D PRS; (b) The plot of the association tests of all T1D patients compared to all controls.

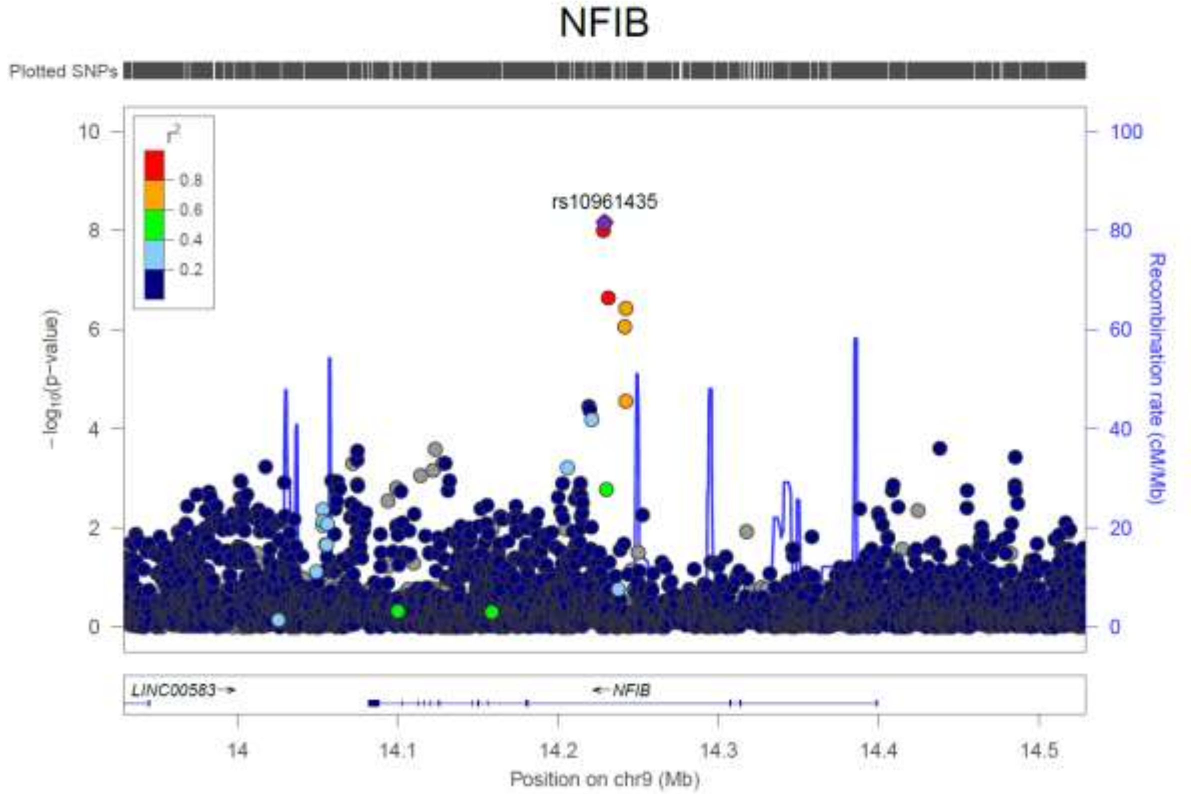


a.

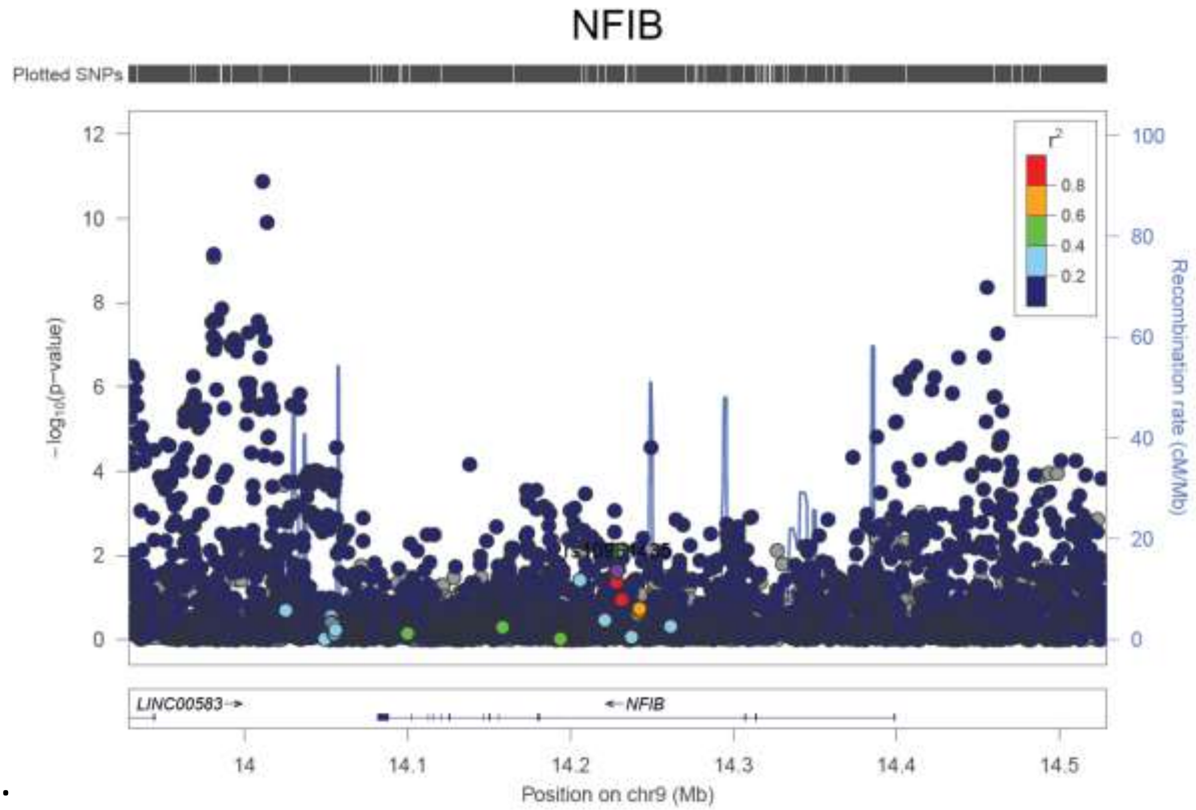


b.

**Supplementary Figure 3.** The LocusZoom plots for each of the *NFIB* locus. (a) The plot of the association tests of T1D patients with low T1D PRS compared to controls with low T1D PRS; (b) The plot of the association tests of all T1D patients compared to all controls.

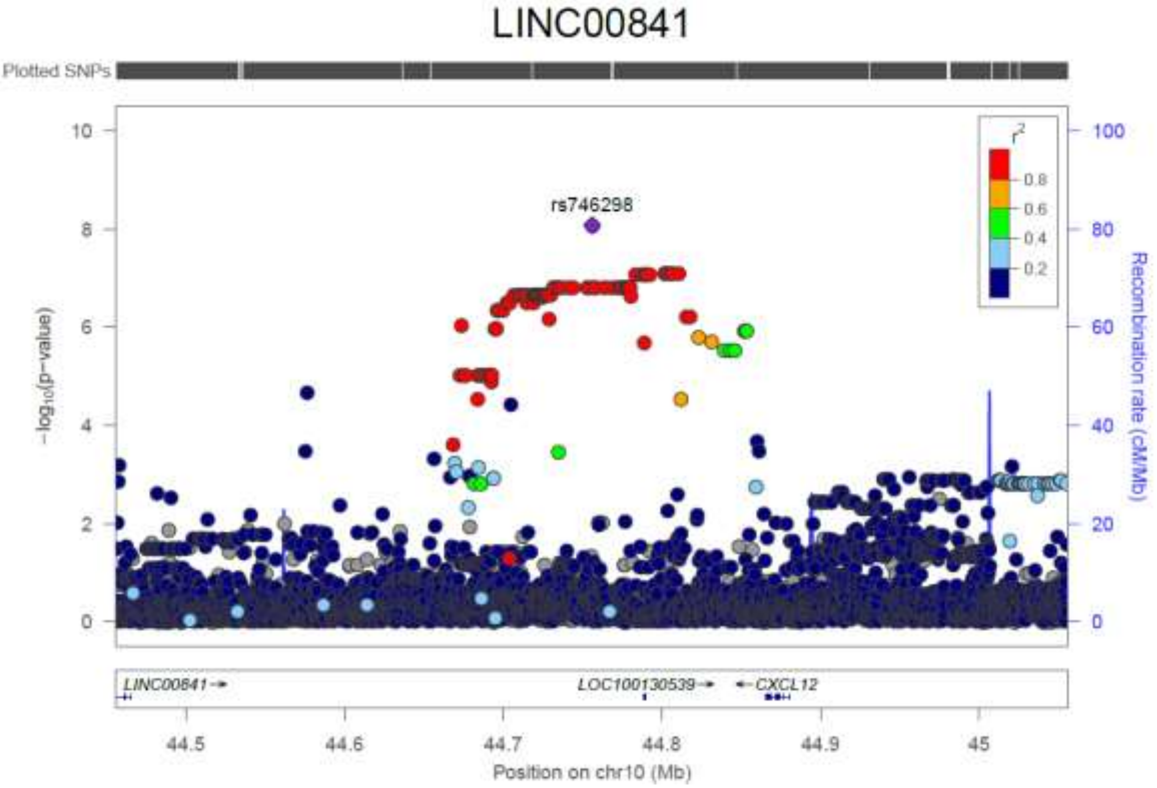


a.

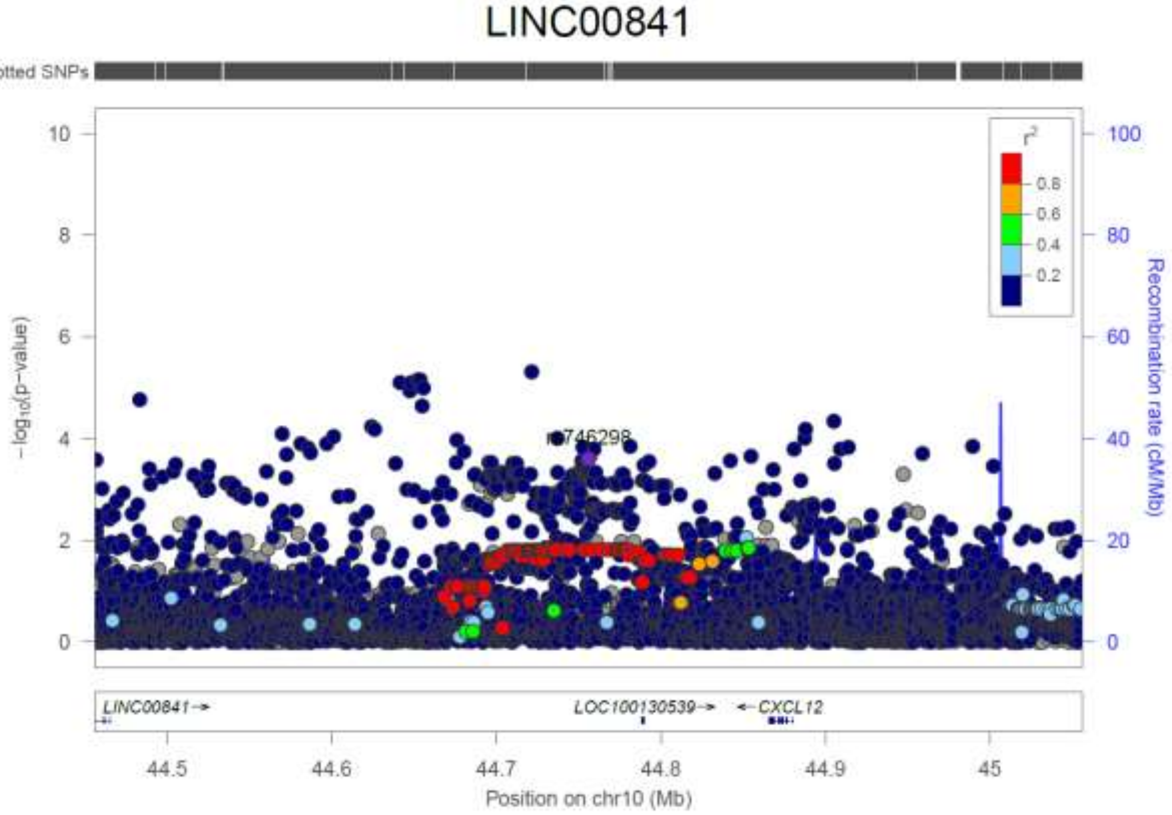


b.

**Supplementary Figure 4.** The LocusZoom plots for each of the *LINC00841/C10orf142* locus. (a) The plot of the association tests of T1D patients with low T1D PRS compared to controls with low T1D PRS; (b) The plot of the association tests of all T1D patients compared to all controls.

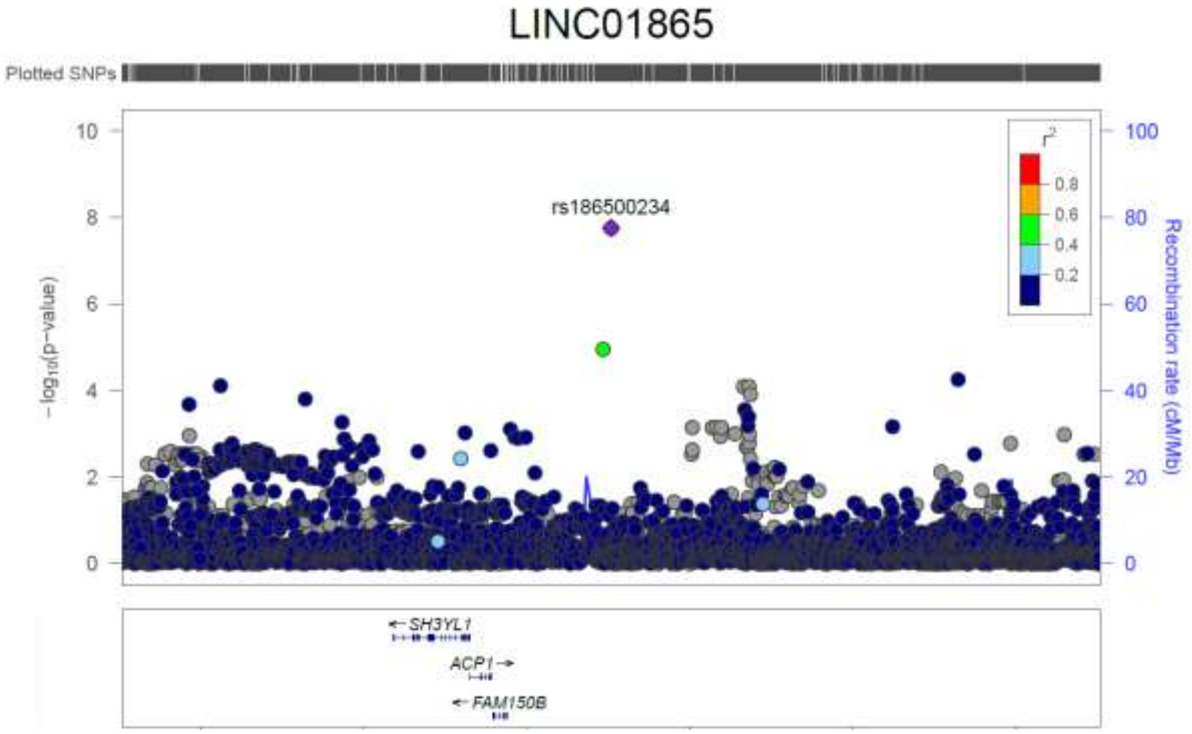


a.

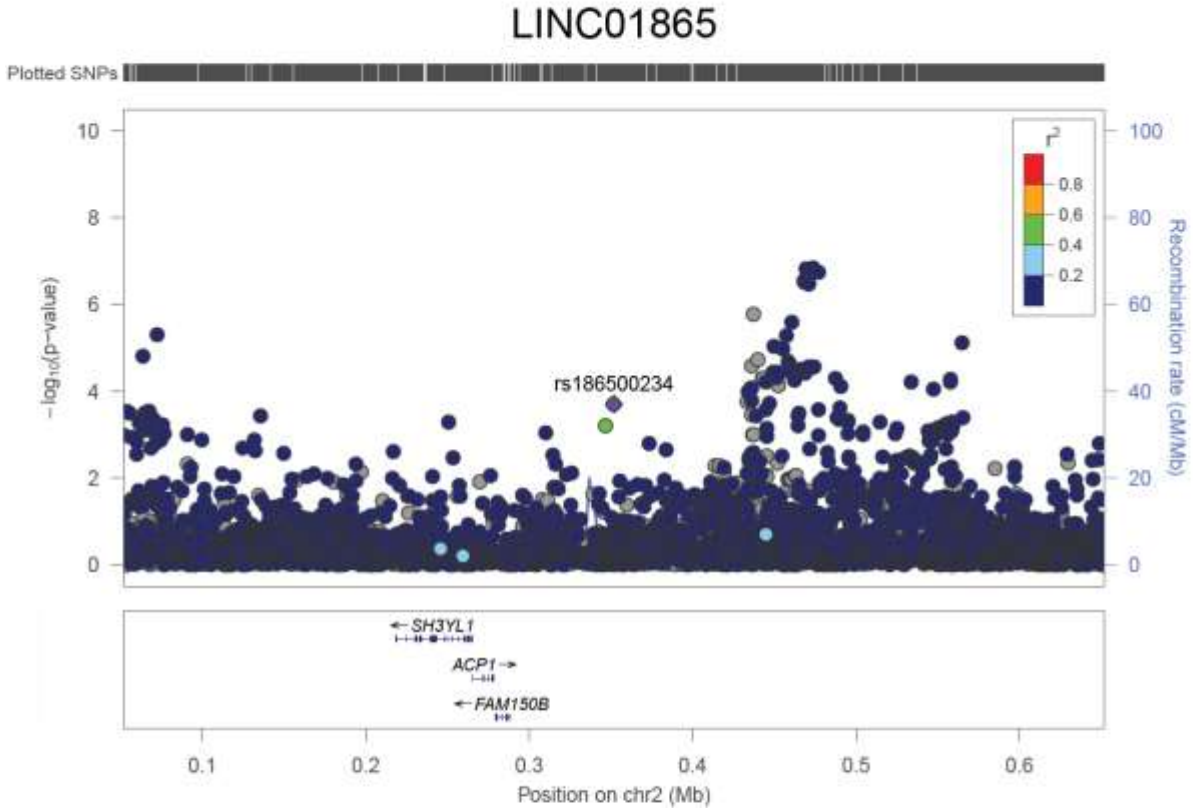


b.

**Supplementary Figure 5.** The LocusZoom plots for each of the *LINC01865/LINC01874* locus. (a) The plot of the association tests of T1D patients with low T1D PRS compared to controls with low T1D PRS; (b) The plot of the association tests of all T1D patients compared to all controls.

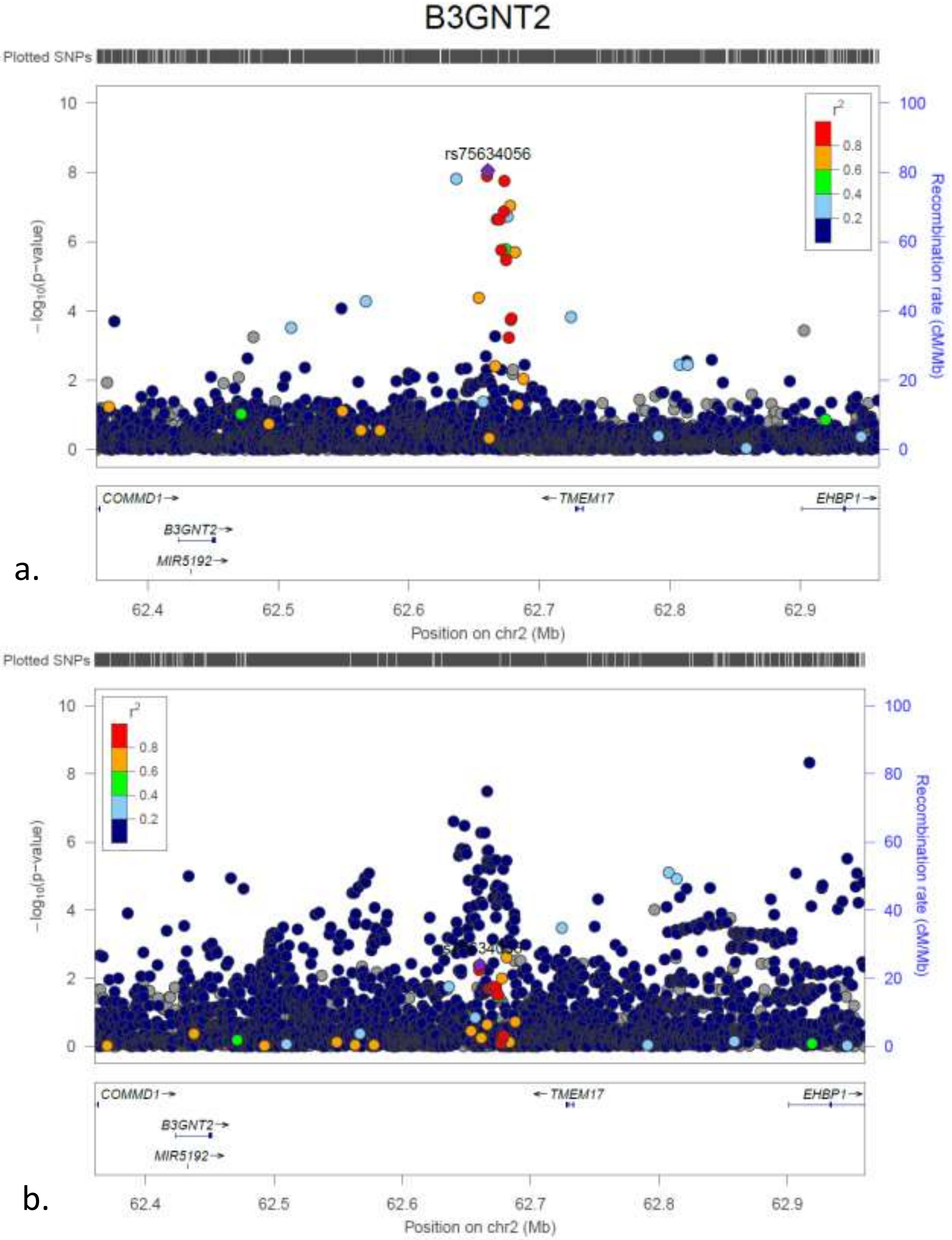


**Supplementary Figure 6.** The LocusZoom plots for each of the *B3GNT2/TMEM17* locus. (a) The plot of the association tests of T1D patients with low T1D PRS compared to controls with low T1D PRS; (b) The plot of the association tests of all T1D patients compared to all controls.

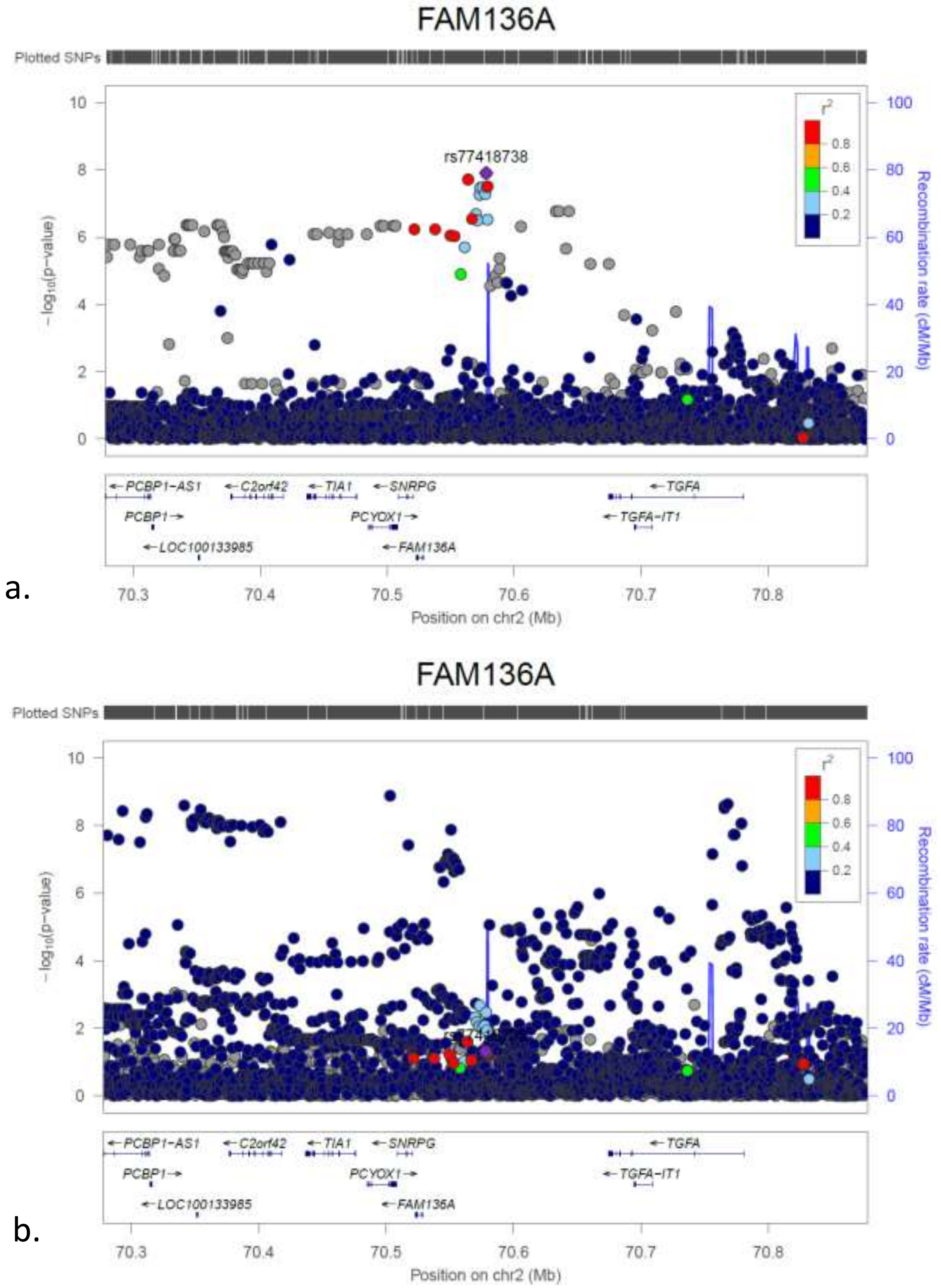


**Supplementary Figure 7.** The LocusZoom plots for each of the *FAM136A/TGFA* locus. (a) The plot of the association tests of T1D patients with low T1D PRS compared to controls with low T1D PRS; (b) The plot of the association tests of all T1D patients compared to all controls.

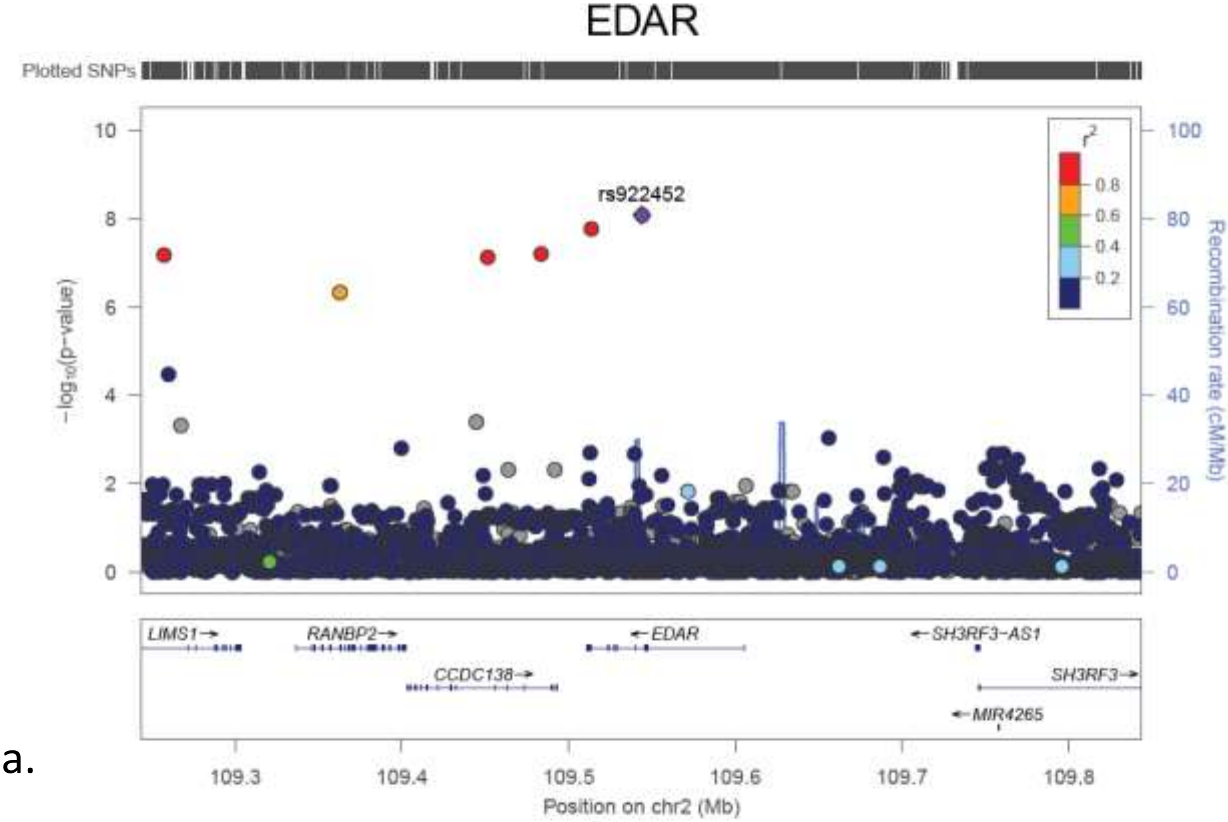


a.

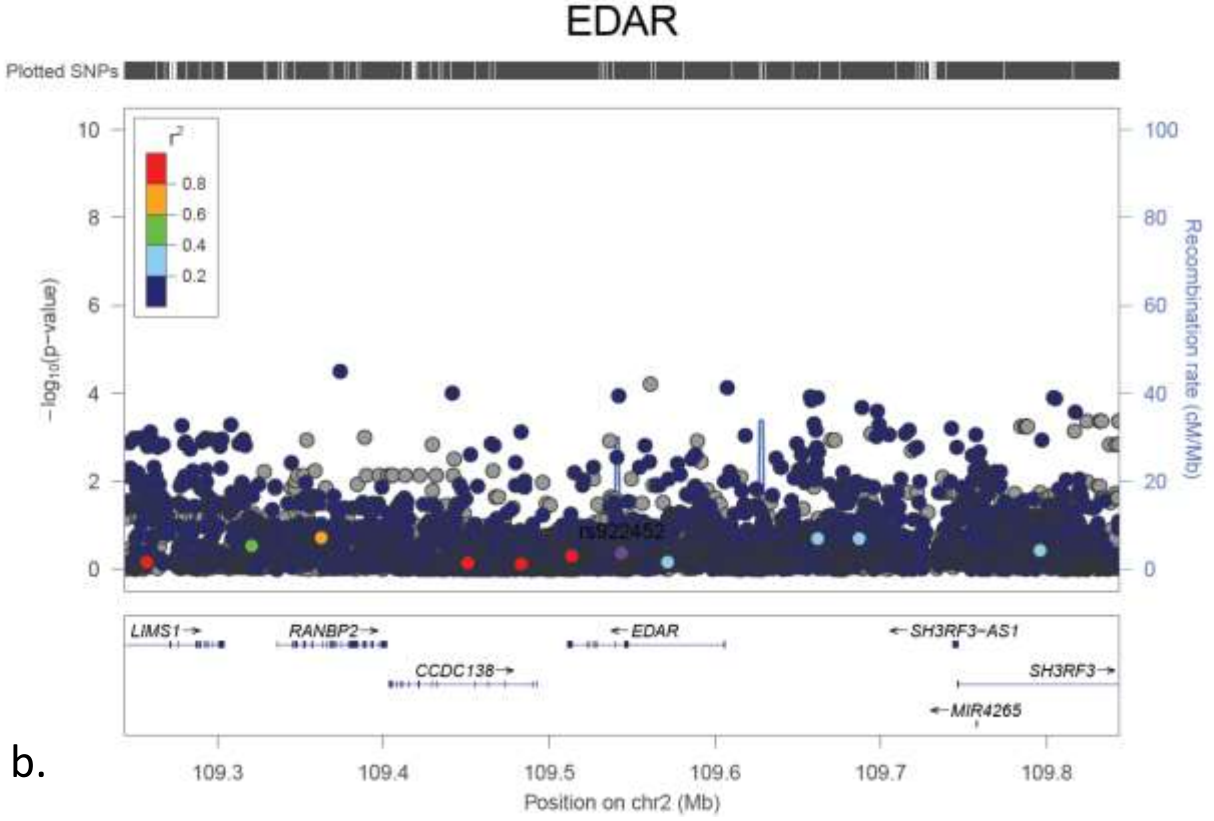


b.

**Supplementary Figure 8.** The LocusZoom plots for each of the *GCC2/EDR* locus. (a) The plot of the association tests of T1D patients with low T1D PRS compared to controls with low T1D PRS; (b) The plot of the association tests of all T1D patients compared to all controls.

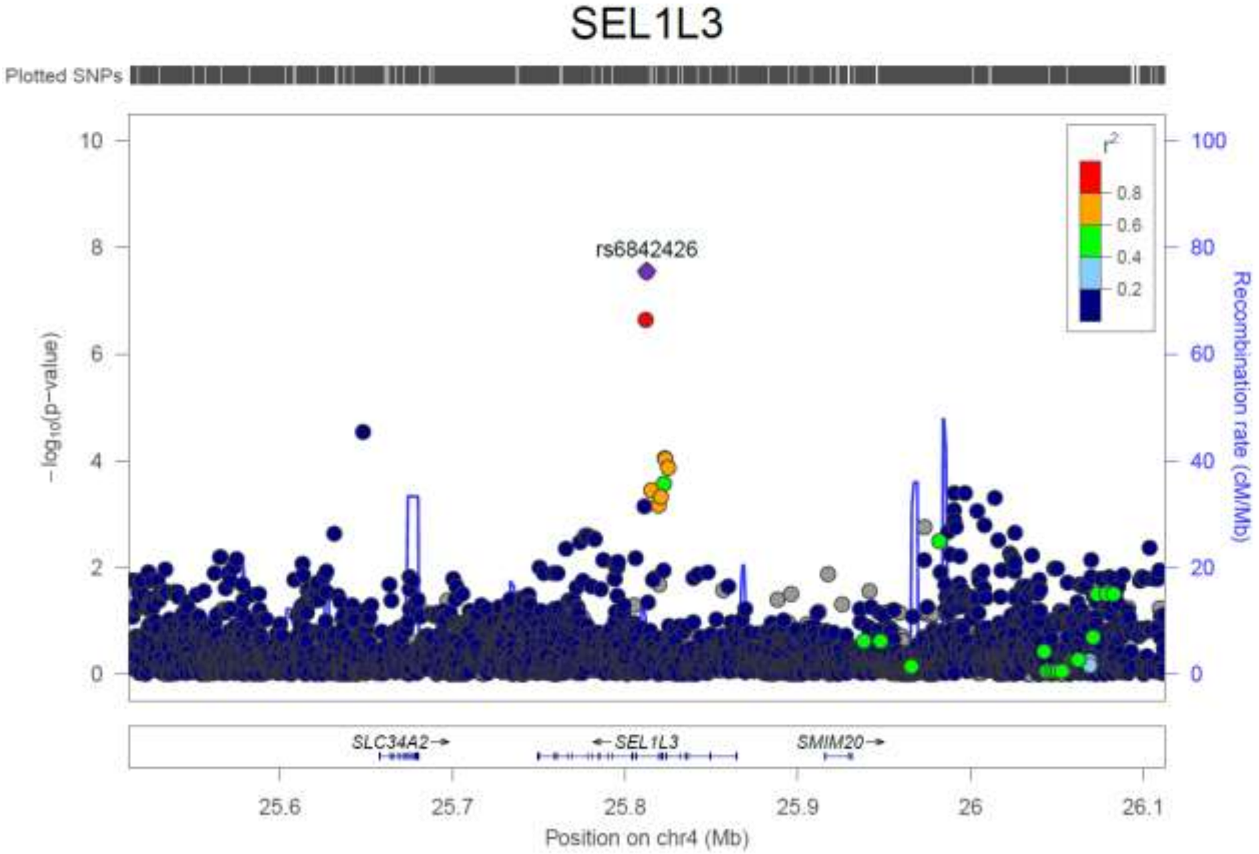


a.

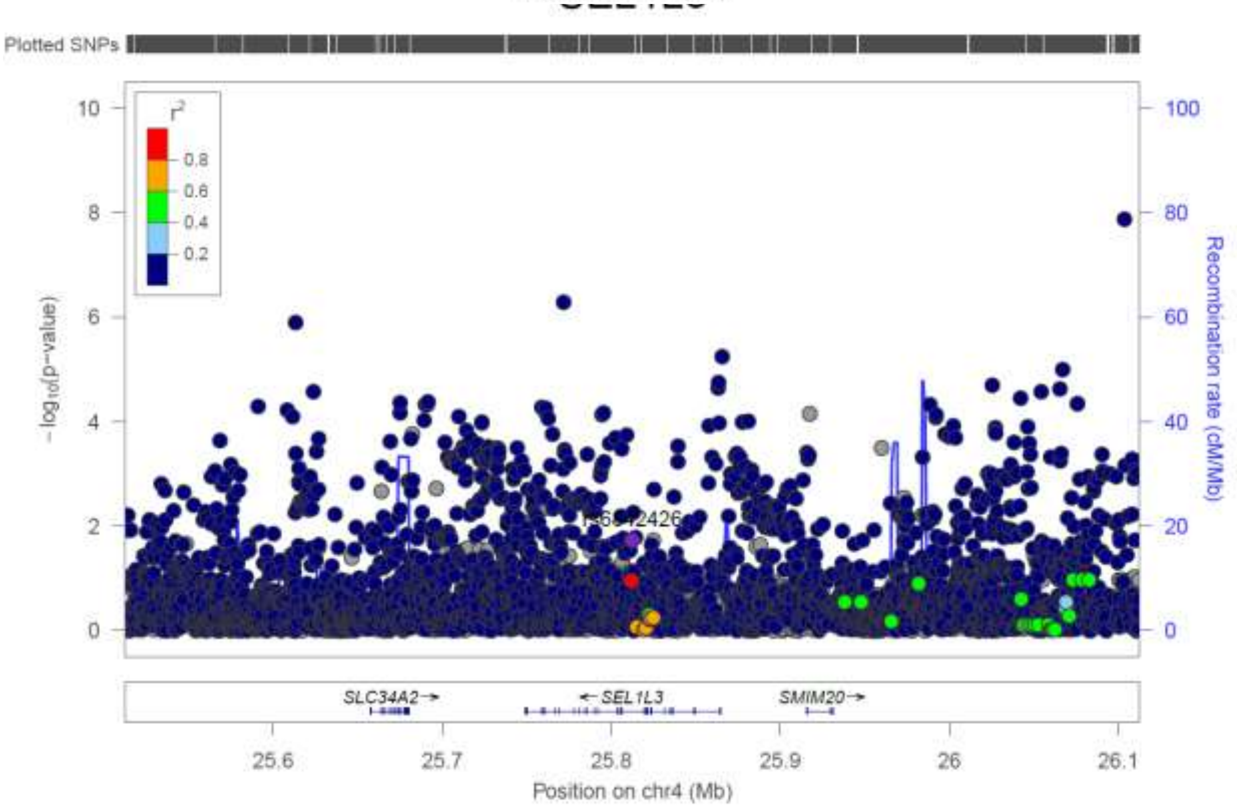


b.

**Supplementary Figure 9.** The LocusZoom plots for each of the *SEL1L3* locus. (a) The plot of the association tests of T1D patients with low T1D PRS compared to controls with low T1D PRS; (b) The plot of the association tests of all T1D patients compared to all controls.

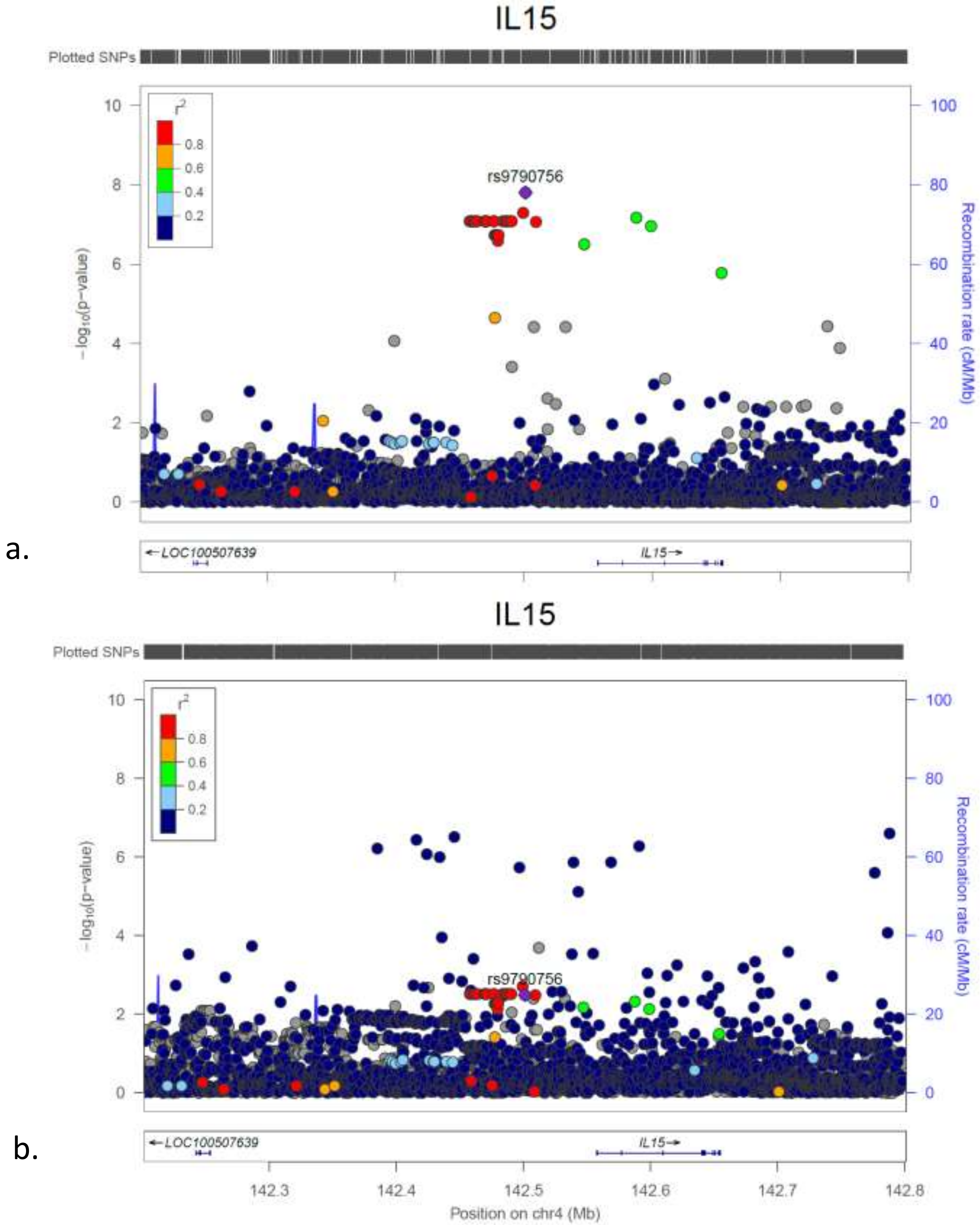


**Supplementary Figure 10.** The LocusZoom plots for each of the *LINC02432/IL15* locus. (a) The plot of the association tests of T1D patients with low T1D PRS compared to controls with low T1D PRS; (b) The plot of the association tests of all T1D patients compared to all controls.

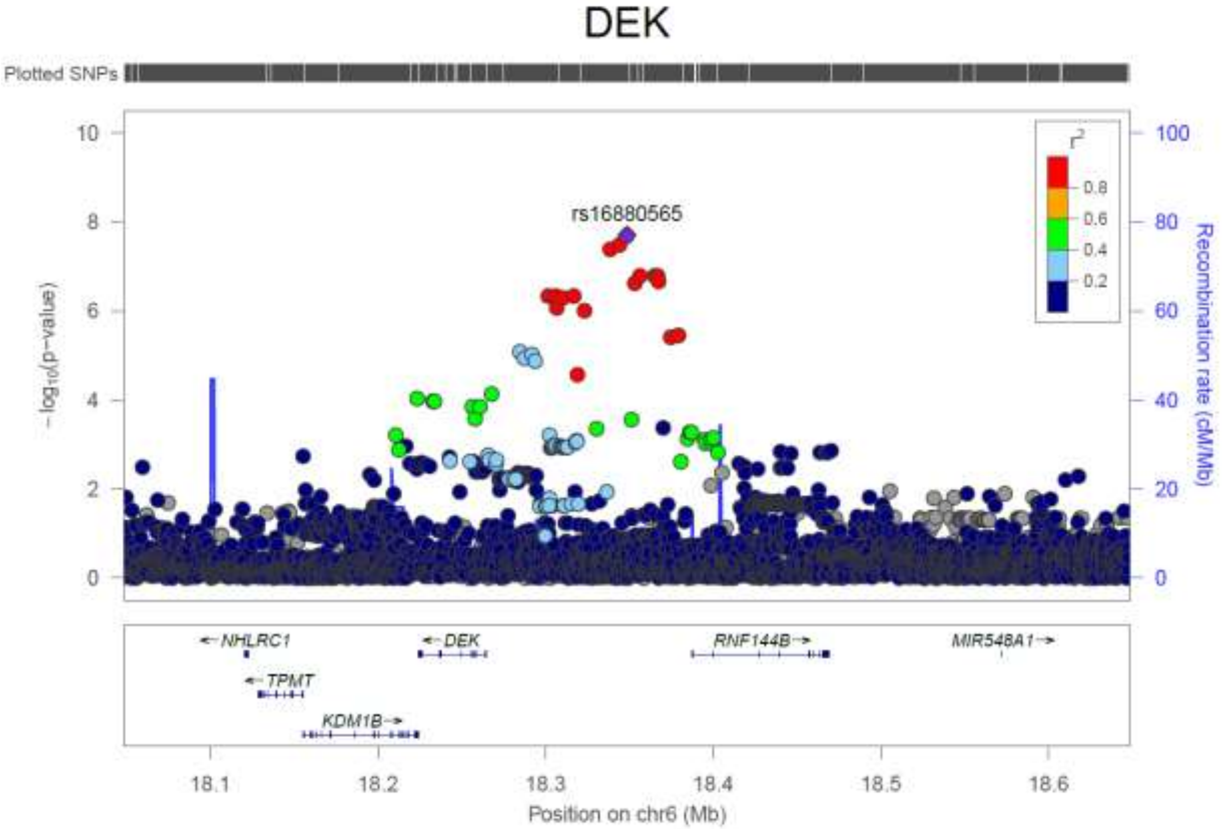


a.

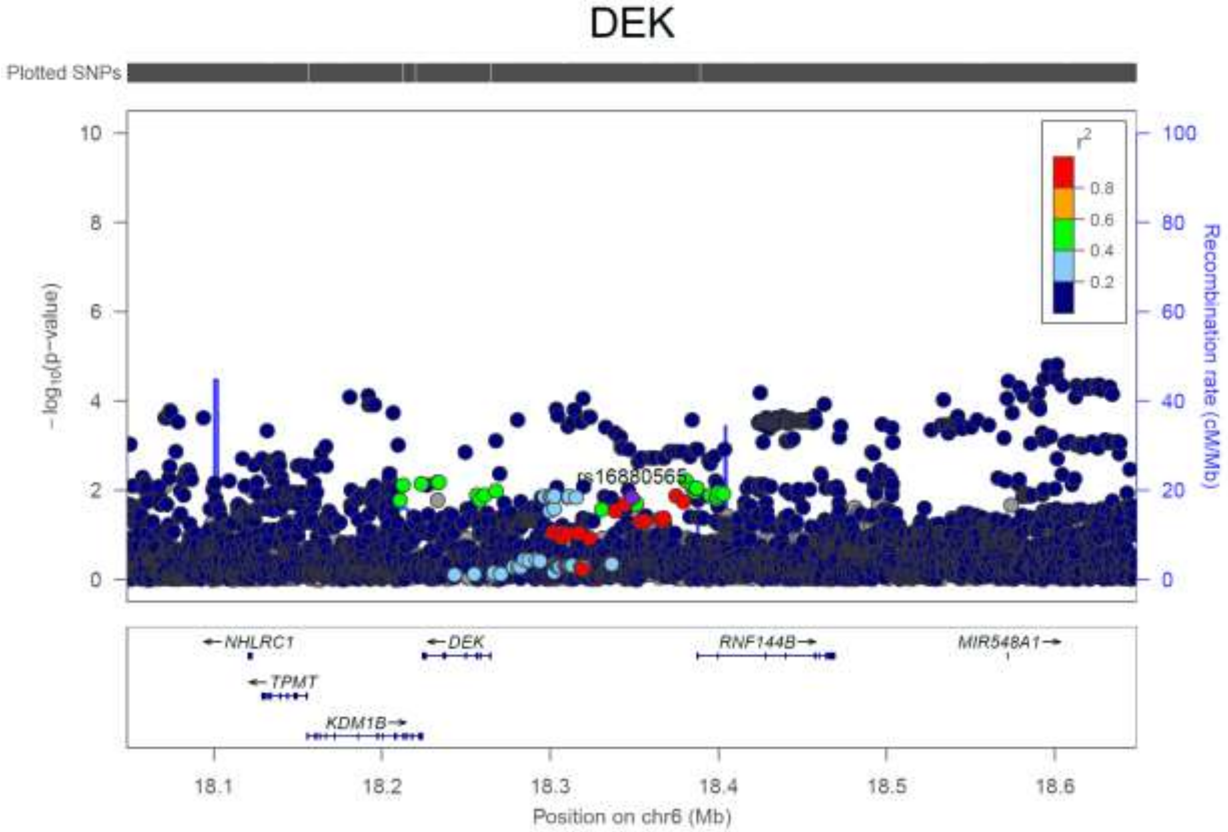


b.

**Supplementary Figure 11.** The LocusZoom plots for each of the *DEK/RNF144B* locus. (a) The plot of the association tests of T1D patients with low T1D PRS compared to controls with low T1D PRS; (b) The plot of the association tests of all T1D patients compared to all controls.

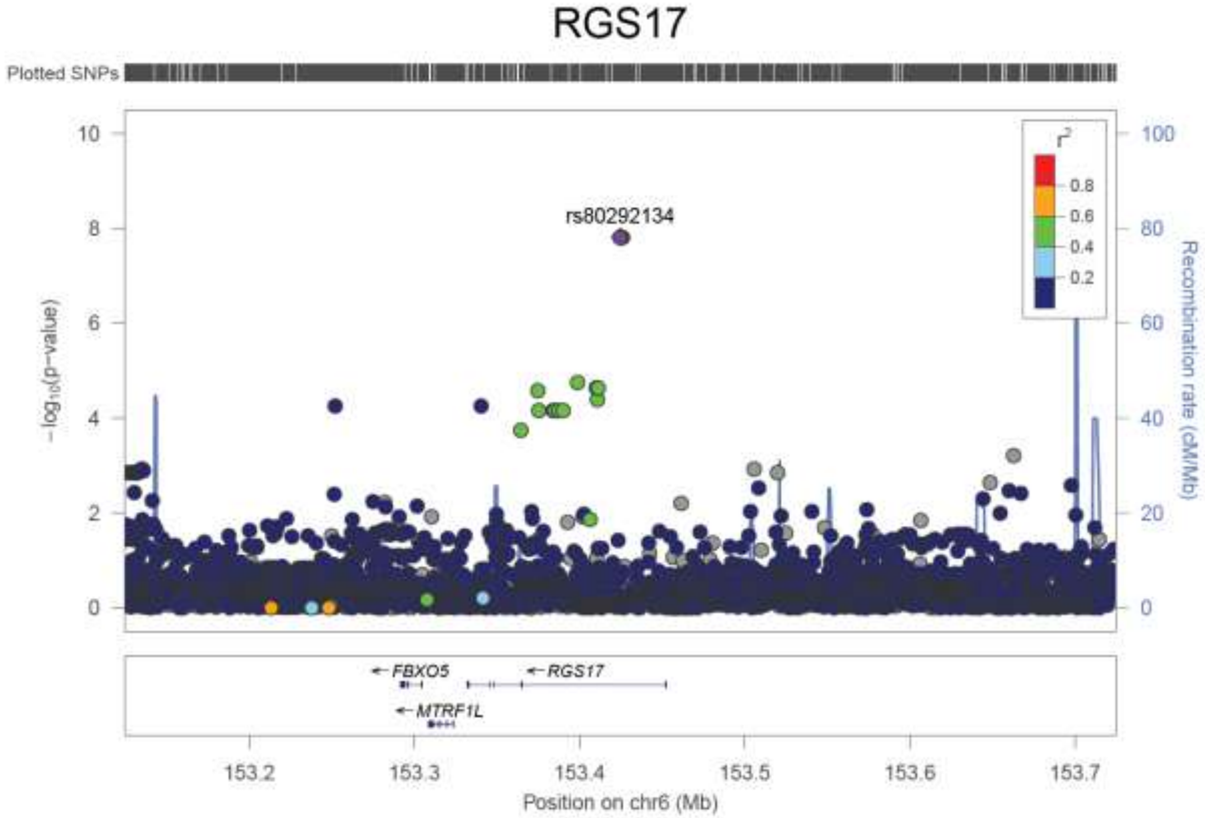


a.

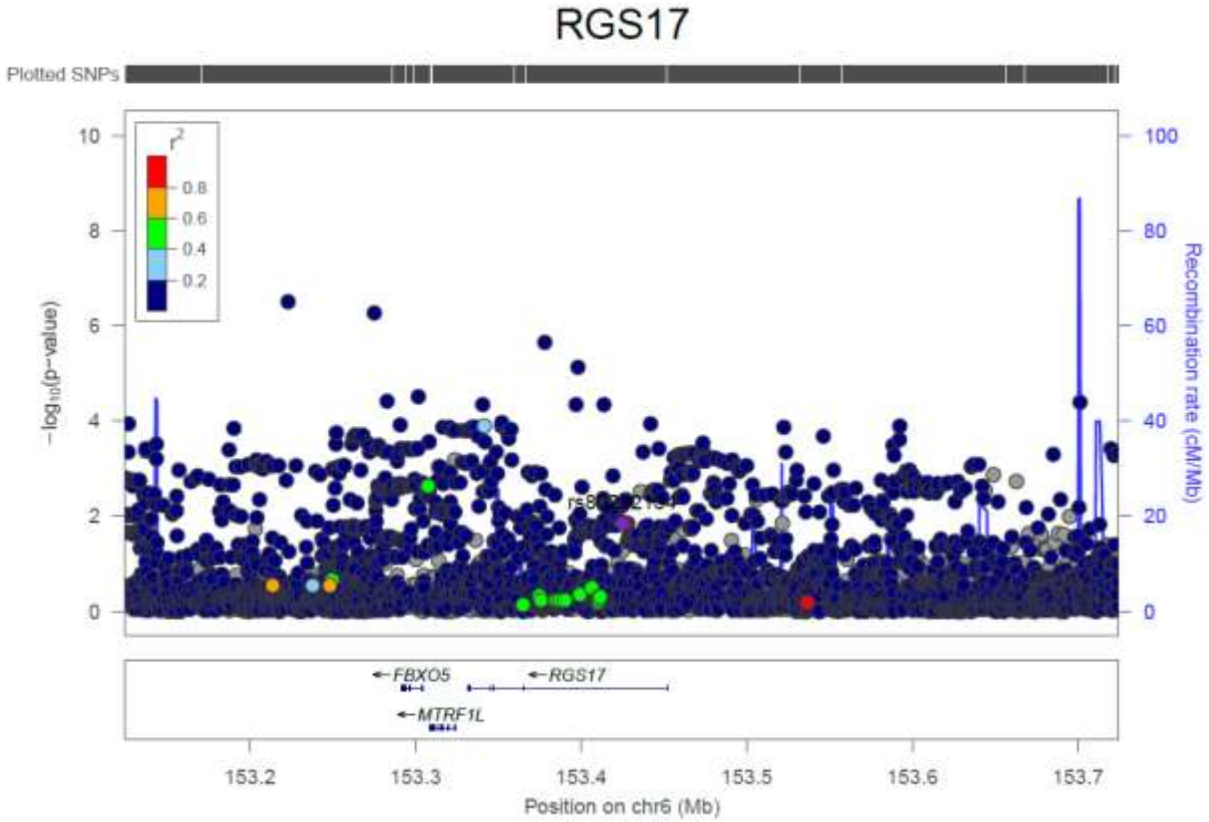


b.

**Supplementary Figure 12.** The LocusZoom plots for each of the *RGS17* locus. (a) The plot of the association tests of T1D patients with low T1D PRS compared to controls with low T1D PRS; (b) The plot of the association tests of all T1D patients compared to all controls.

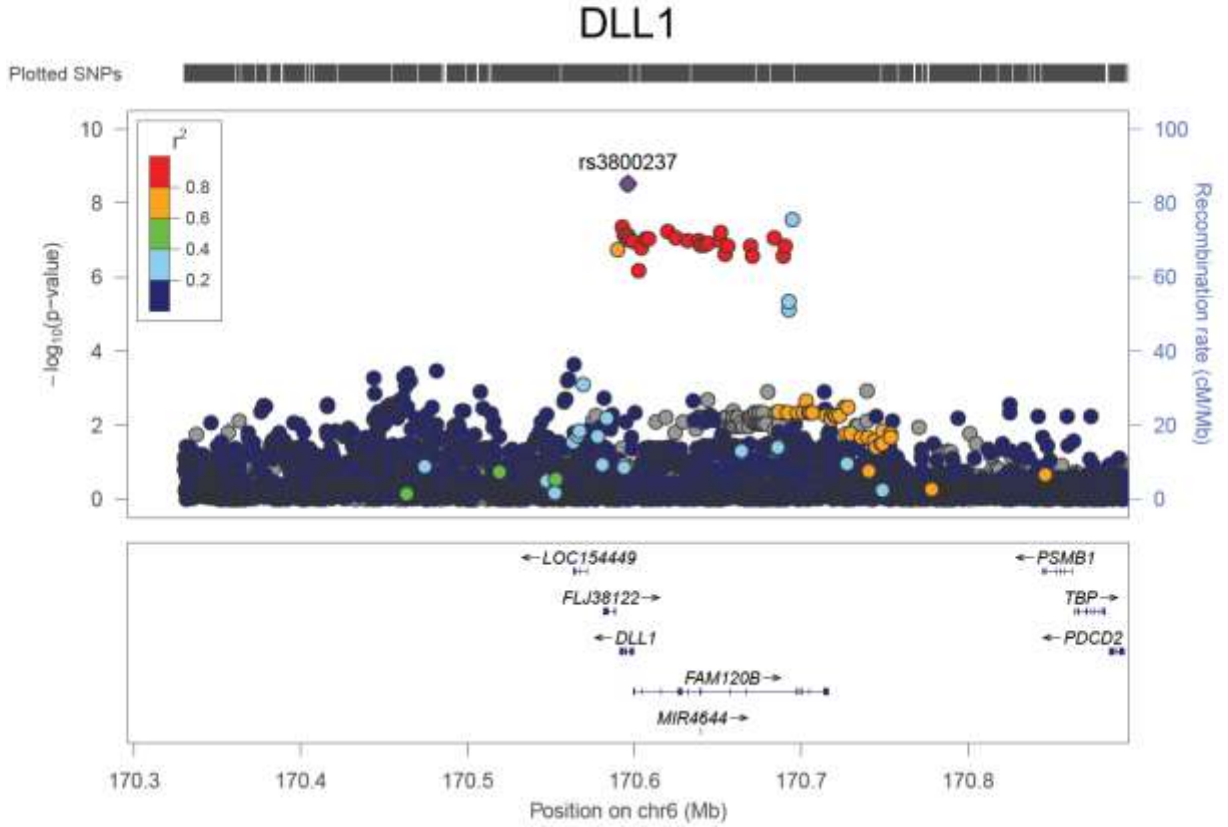


a.

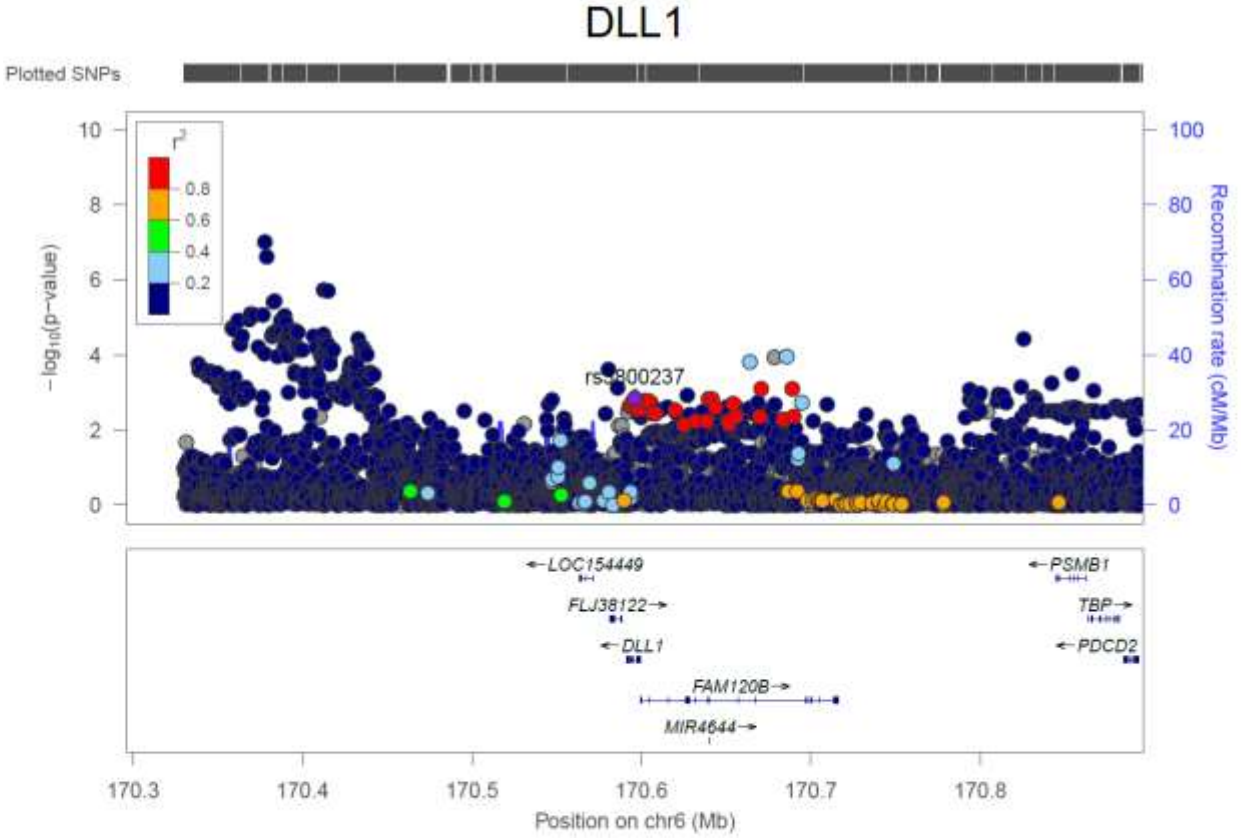


b.

**Supplementary Figure 13.** The LocusZoom plots for each of the *DLL1/FAM120B* locus. (a) The plot of the association tests of T1D patients with low T1D PRS compared to controls with low T1D PRS; (b) The plot of the association tests of all T1D patients compared to all controls.

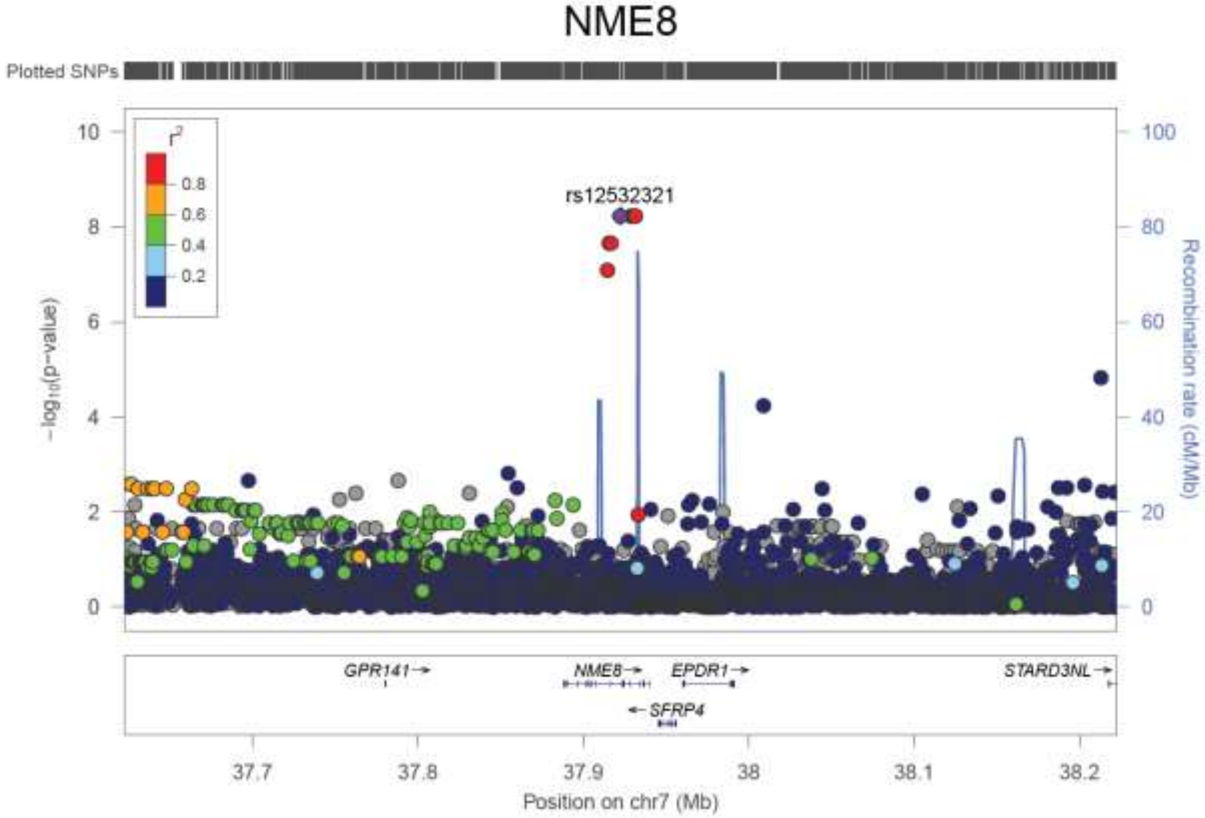


a.

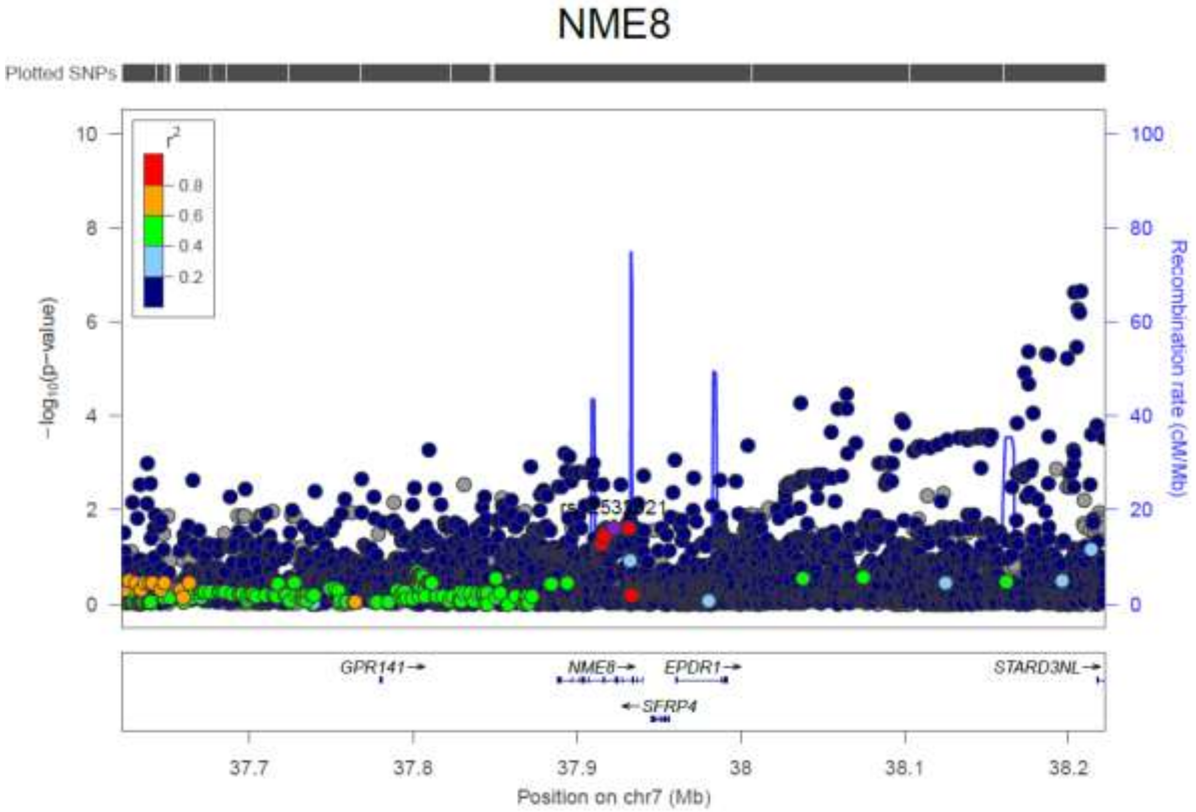


b.

**Supplementary Figure 14.** The LocusZoom plots for each of the *NME8* locus. (a) The plot of the association tests of T1D patients with low T1D PRS compared to controls with low T1D PRS; (b) The plot of the association tests of all T1D patients compared to all controls.

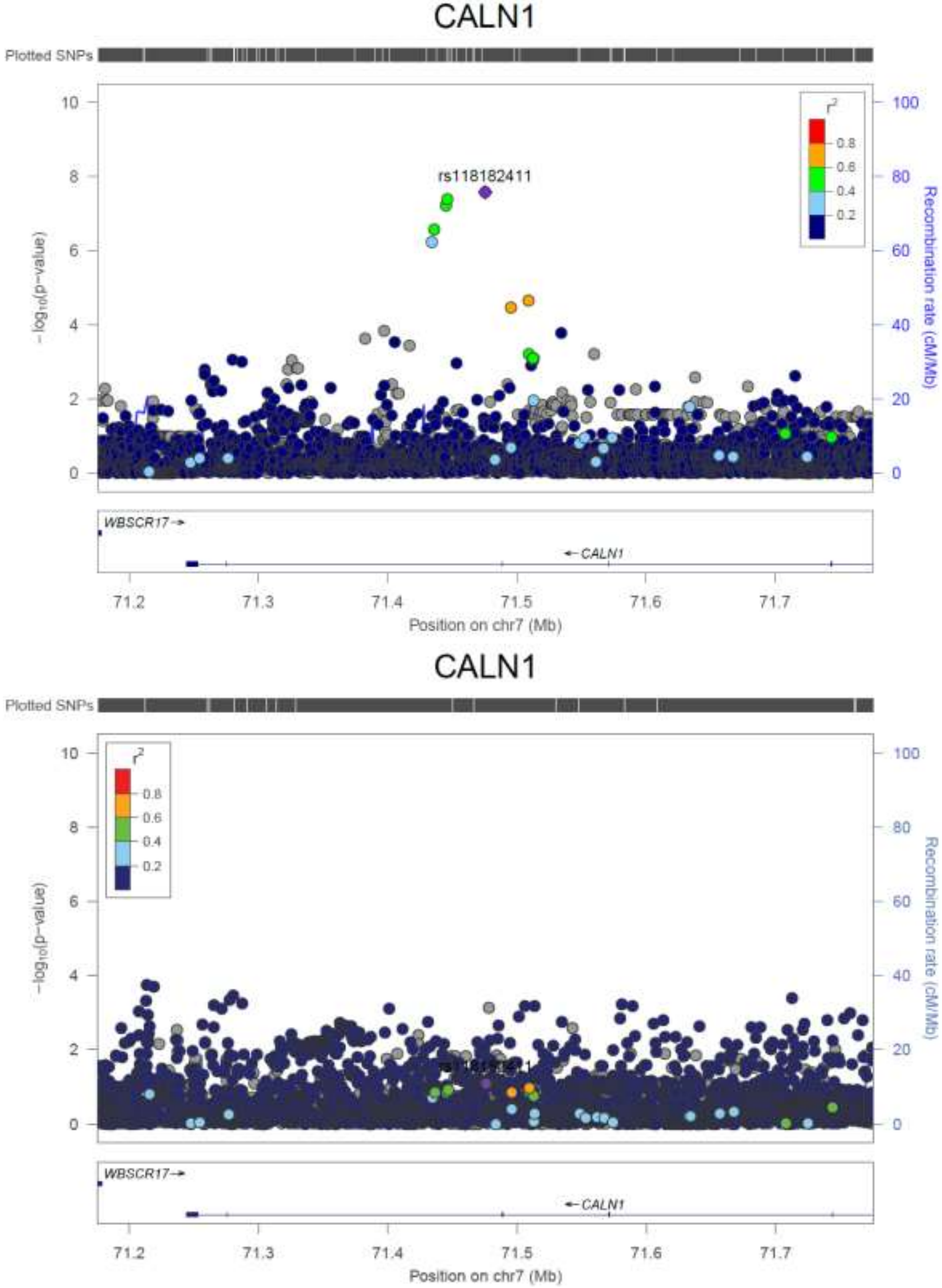


a.

b.

**Supplementary Figure 15.** The LocusZoom plots for each of the *CALN1* locus. (a) The plot of the association tests of T1D patients with low T1D PRS compared to controls with low T1D PRS; (b) The plot of the association tests of all T1D patients compared to all controls.

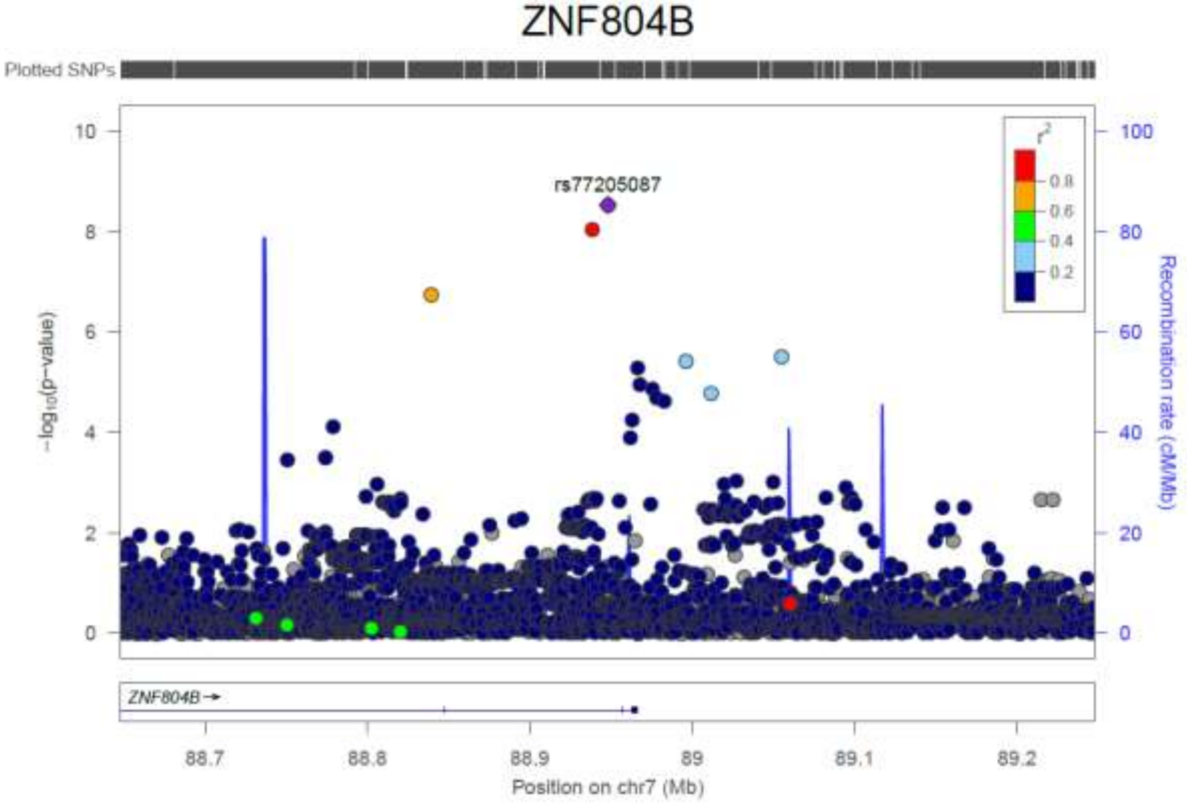


a.

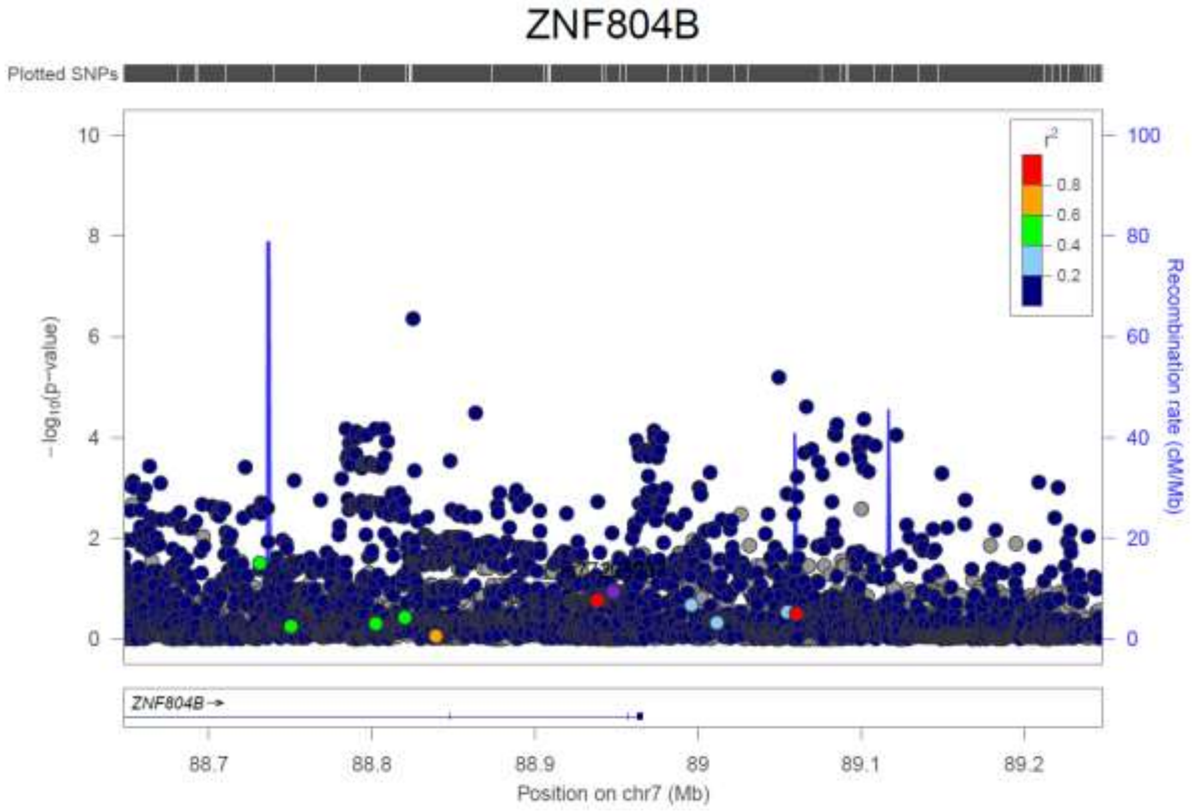


b.

**Supplementary Figure 16.** The LocusZoom plots for each of the *ZNF804B* locus. (a) The plot of the association tests of T1D patients with low T1D PRS compared to controls with low T1D PRS; (b) The plot of the association tests of all T1D patients compared to all controls.

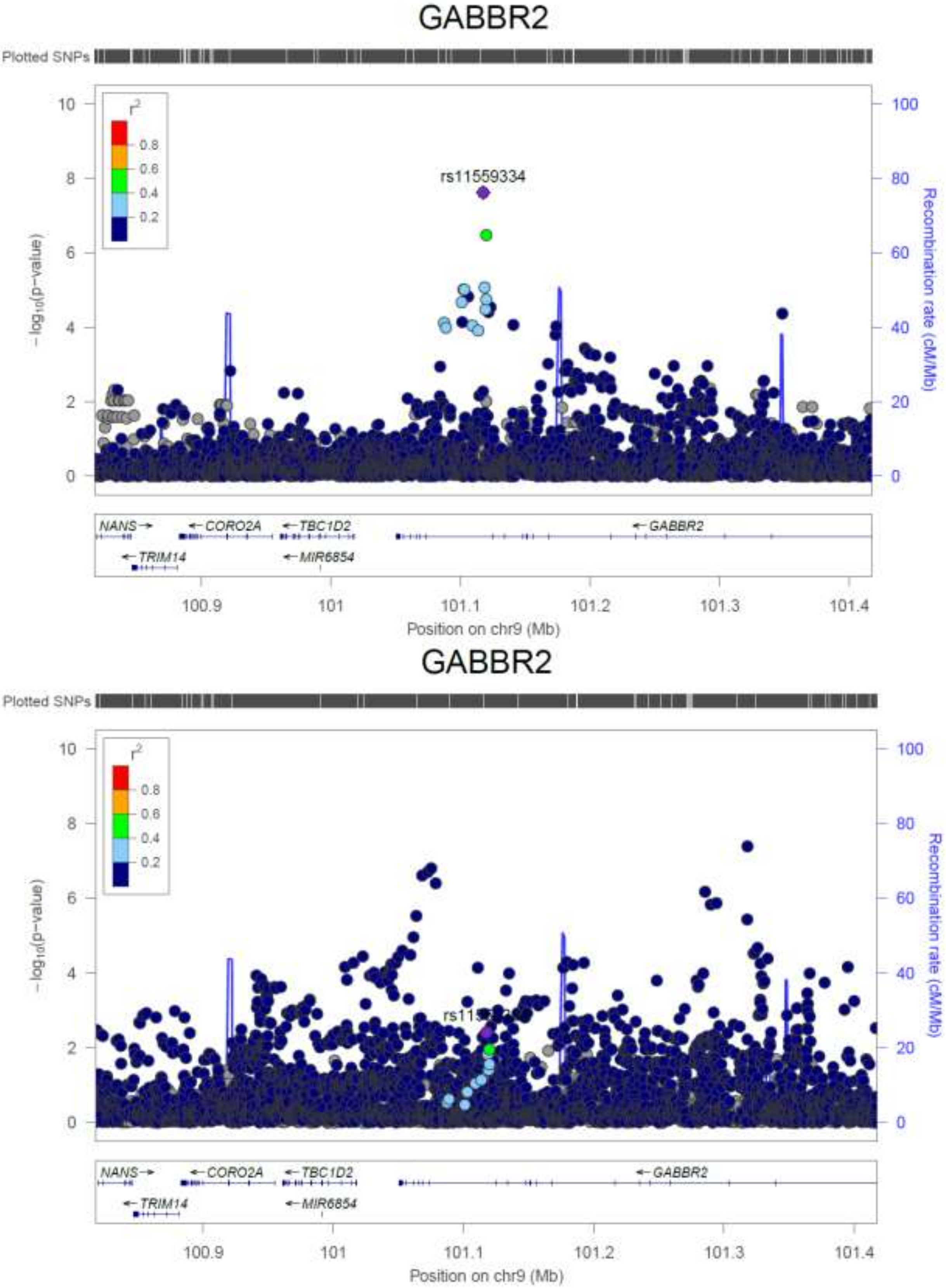


a.

b.

**Supplementary Figure 17.** The LocusZoom plots for each of the *GABBR2* locus. (a) The plot of the association tests of T1D patients with low T1D PRS compared to controls with low T1D PRS; (b) The plot of the association tests of all T1D patients compared to all controls.

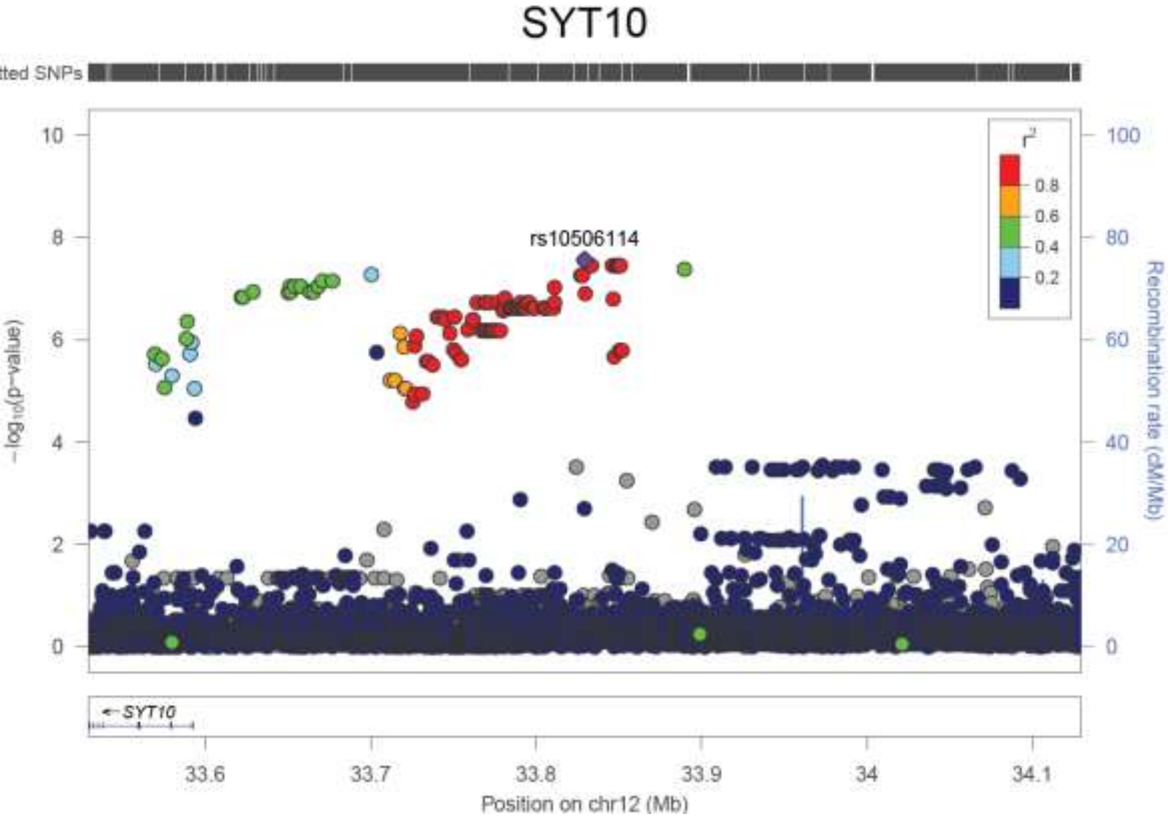


a.

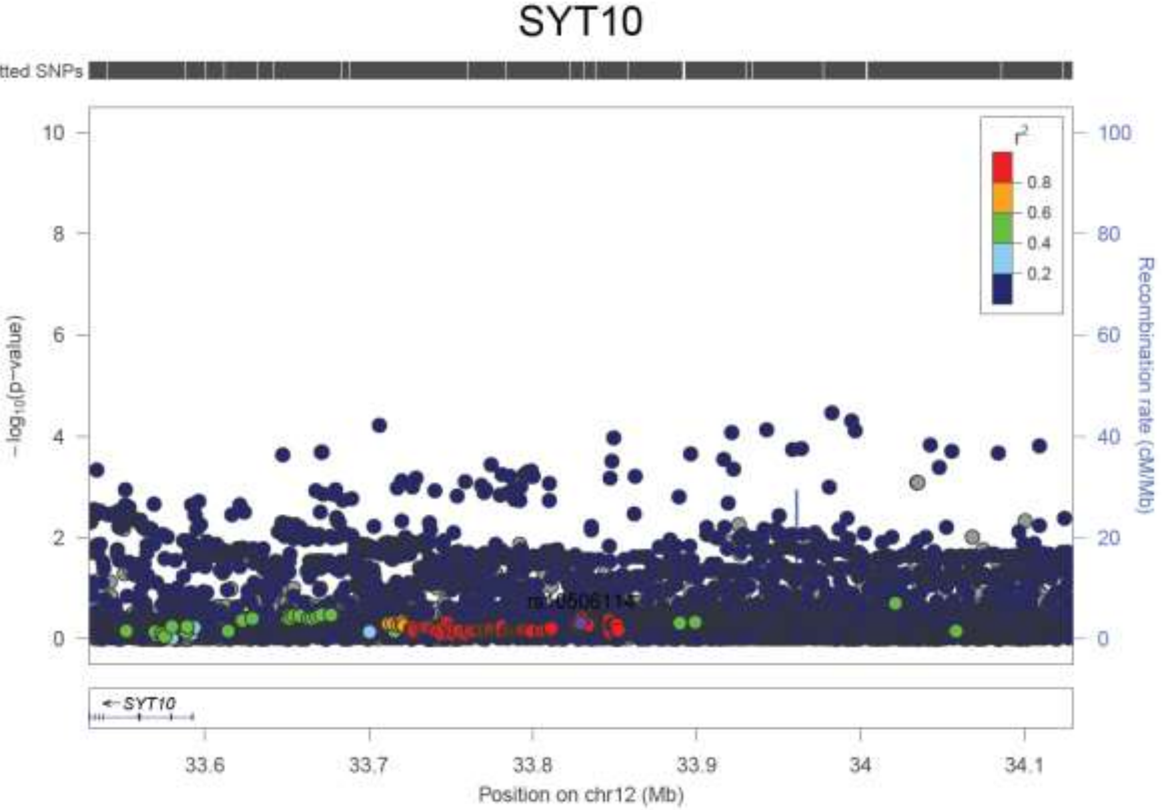


b.

**Supplementary Figure 18.** The LocusZoom plots for each of the *SYT10/ALG10* locus. (a) The plot of the association tests of T1D patients with low T1D PRS compared to controls with low T1D PRS; (b) The plot of the association tests of all T1D patients compared to all controls.

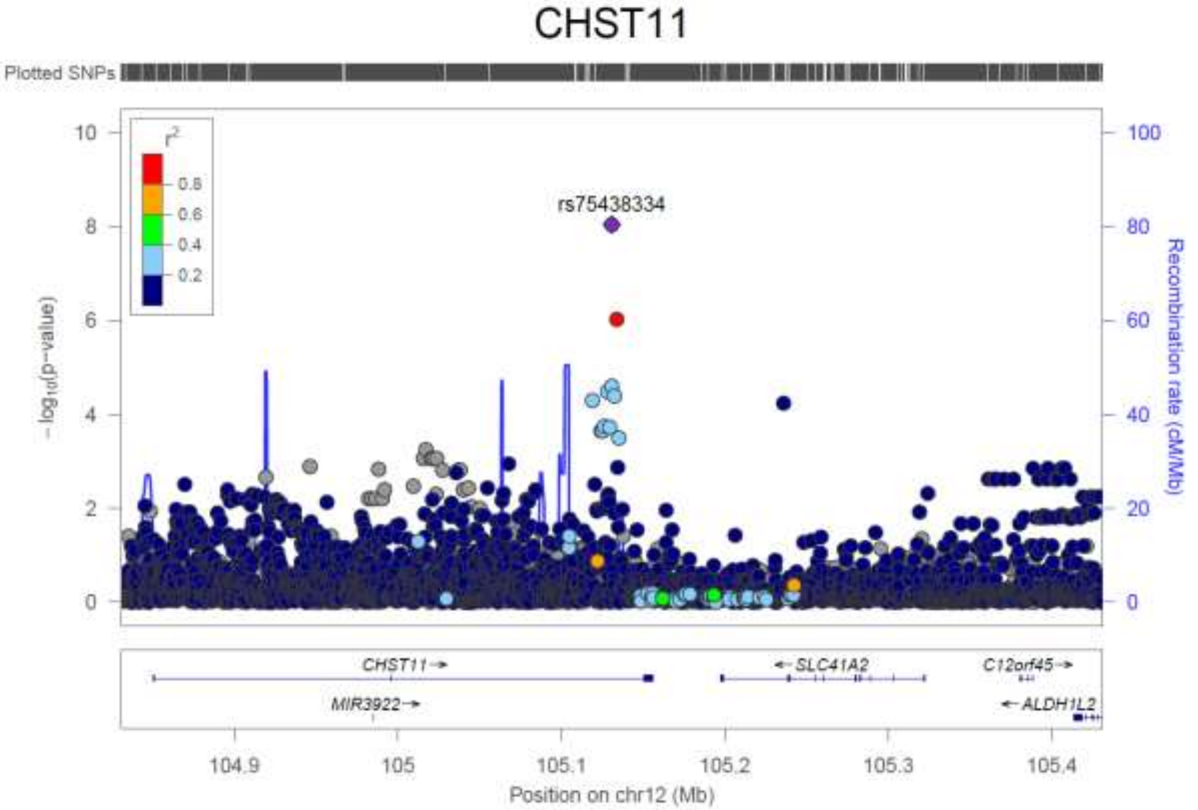


a.

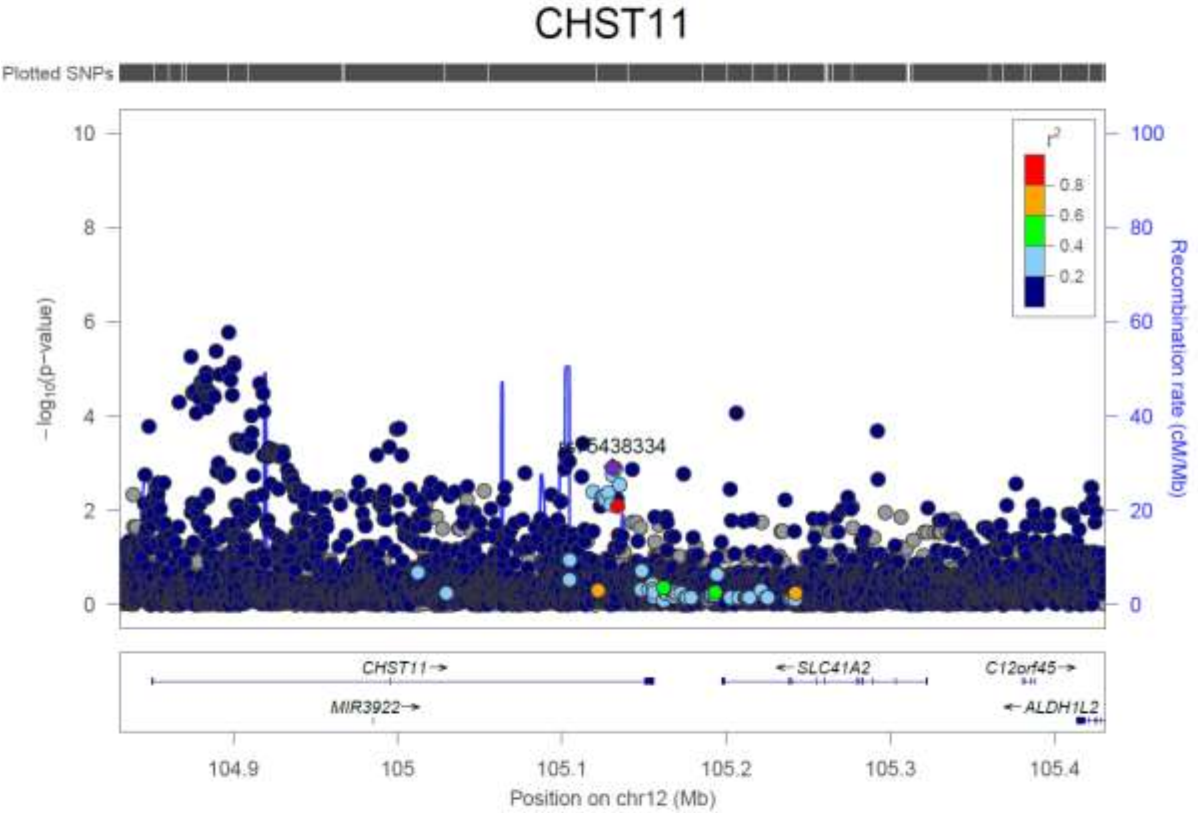


b.

**Supplementary Figure 19.** The LocusZoom plots for each of the *CHST11* locus. (a) The plot of the association tests of T1D patients with low T1D PRS compared to controls with low T1D PRS; (b) The plot of the association tests of all T1D patients compared to all controls.

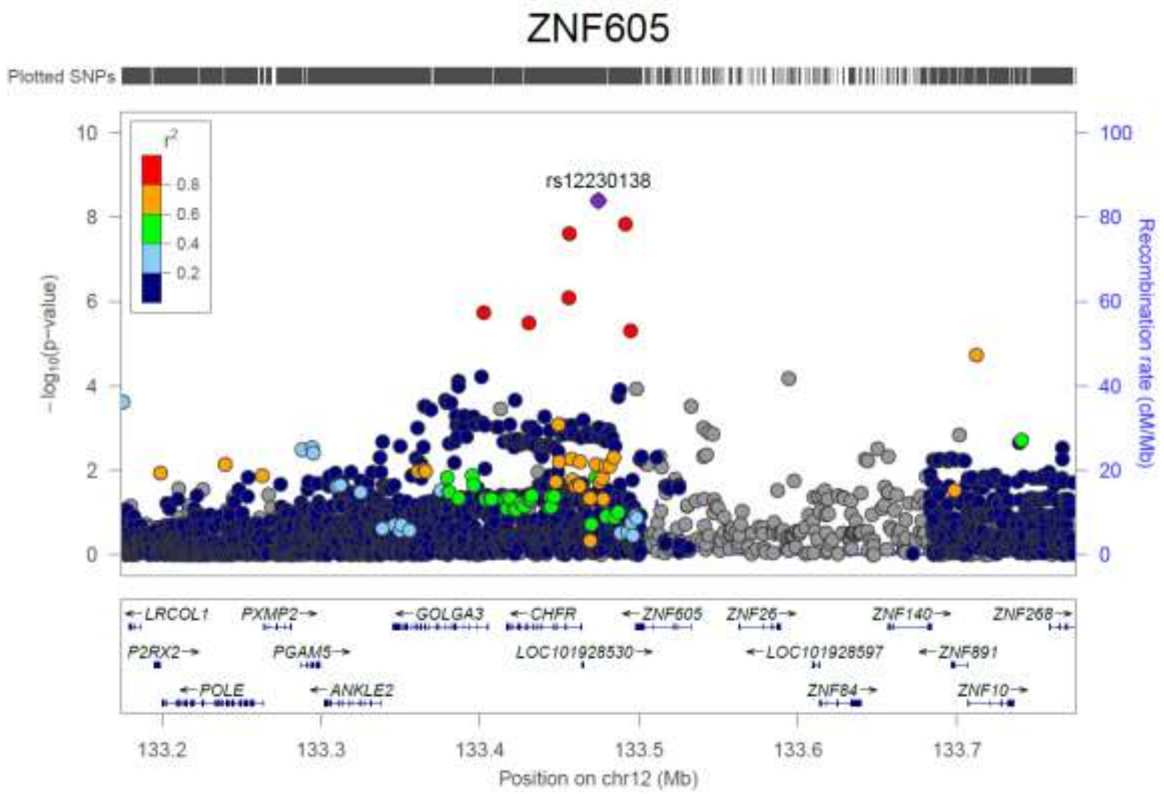


a.

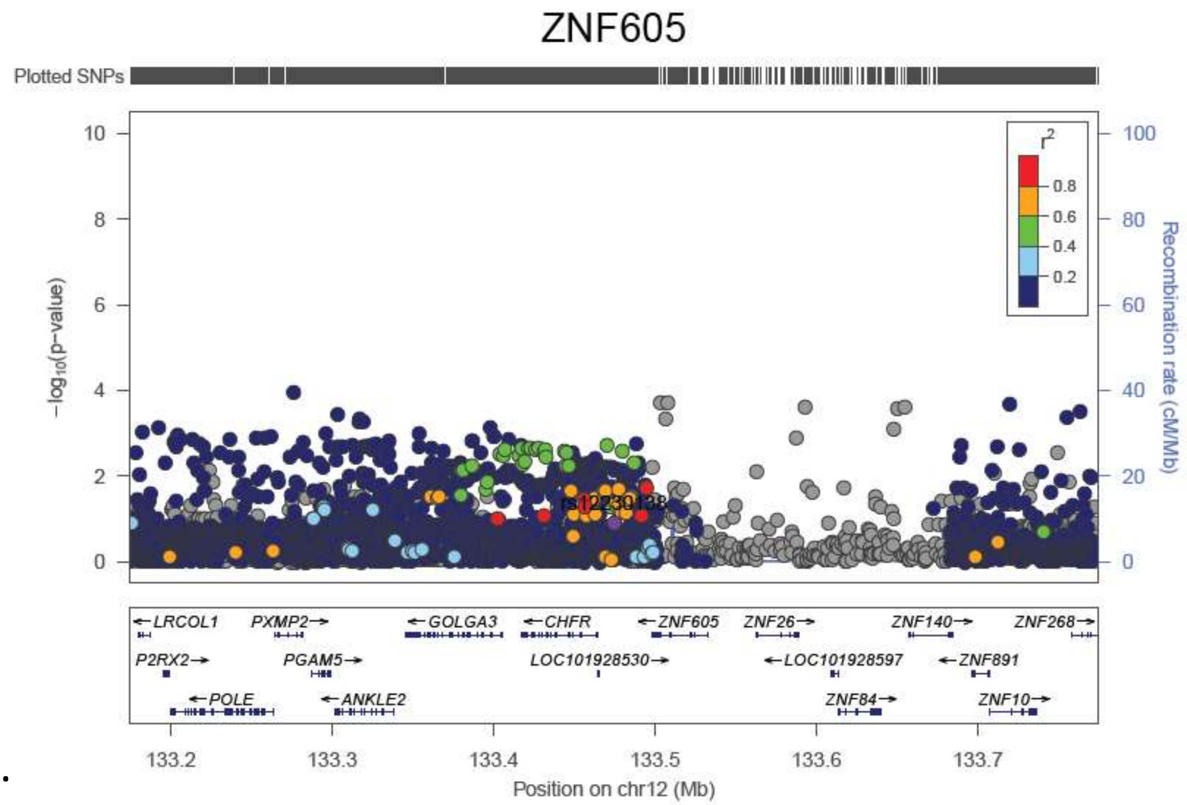


b.

**Supplementary Figure 20.** The LocusZoom plots for each of the *CHFR/LOC101928530/ZNF605* locus. (a) The plot of the association tests of T1D patients with low T1D PRS compared to controls with low T1D PRS; (b) The plot of the association tests of all T1D patients compared to all controls.

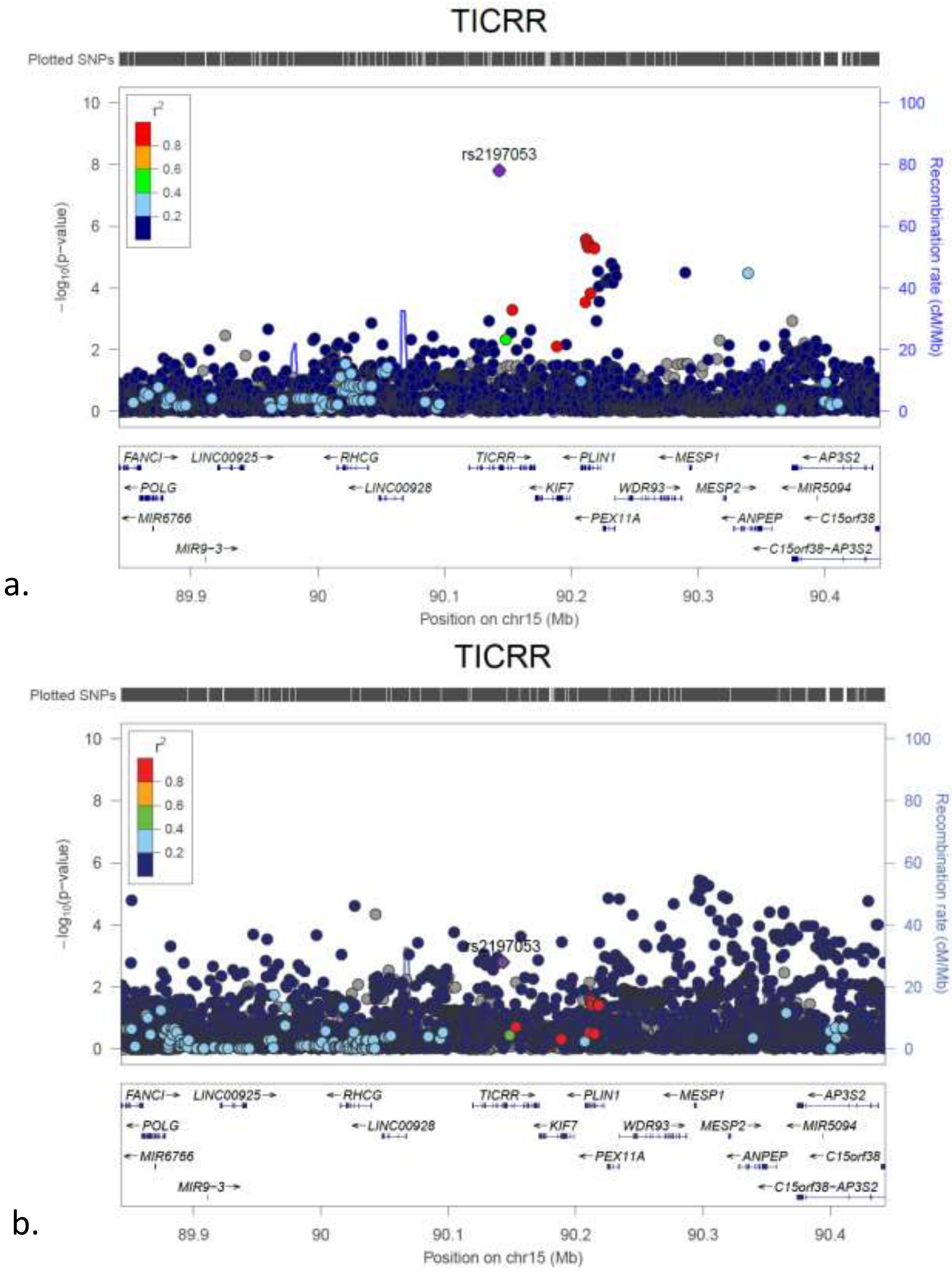


**Supplementary Figure 21.** The LocusZoom plots for each of the *TICRR* locus. (a) The plot of the association tests of T1D patients with low T1D PRS compared to controls with low T1D PRS; (b) The plot of the association tests of all T1D patients compared to all controls.

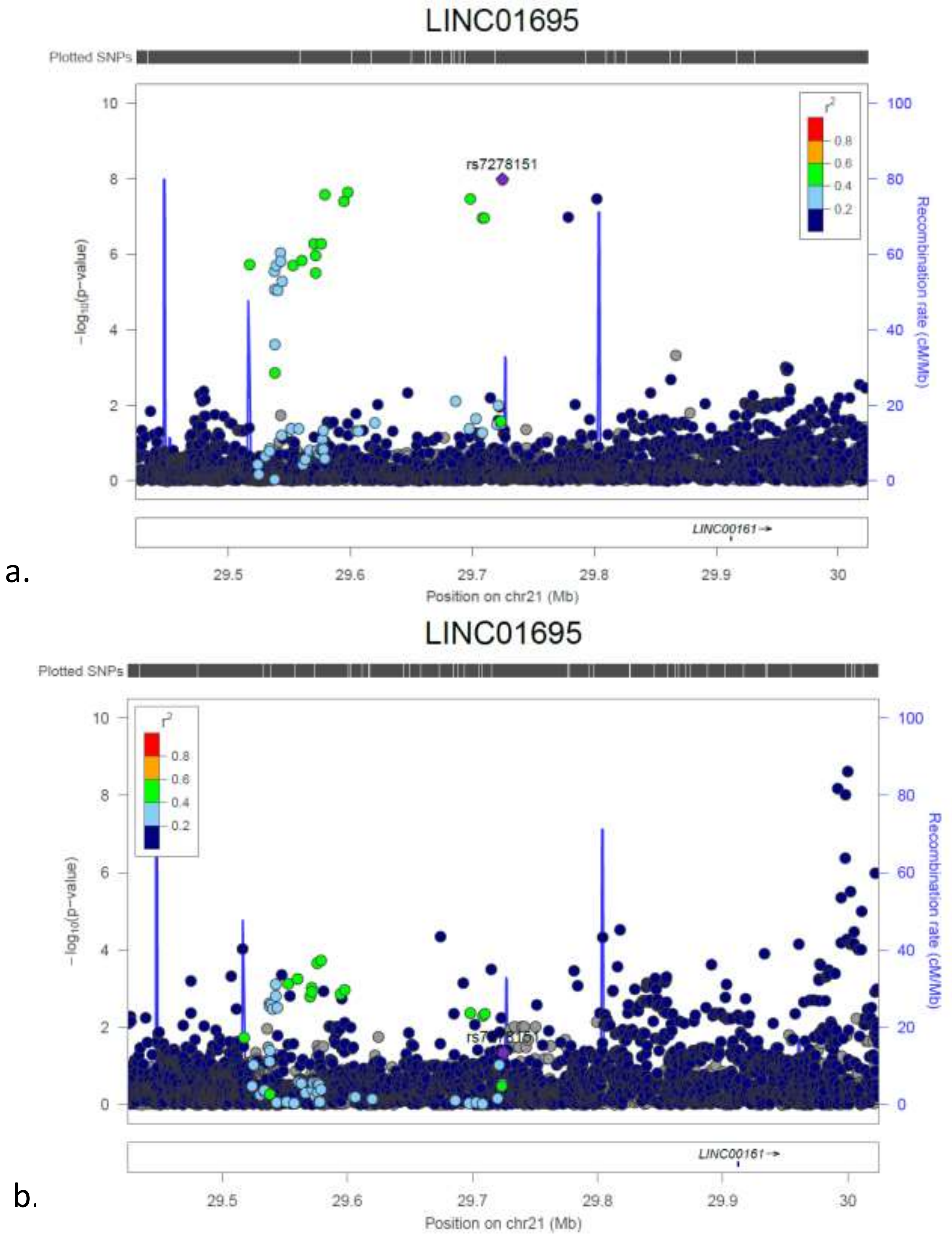


**Supplementary Figure 22.** The LocusZoom plots for each of the *LINC01695/LINC00161* locus. (a) The plot of the association tests of T1D patients with low T1D PRS compared to controls with low T1D PRS; (b) The plot of the association tests of all T1D patients compared to all controls.
